# How Food Environment Impacts Dietary Consumption and Body Weight: A Country-wide Observational Study of 2.3 Billion Food Logs

**DOI:** 10.1101/2020.09.29.20204099

**Authors:** Tim Althoff, Hamed Nilforoshan, Jenna Hua, Jure Leskovec

**Affiliations:** Allen School of Computer Science & Engineering, University of Washington; Department of Computer Science, Stanford University; Stanford Prevention Research Center, Department of Medicine, Stanford University School of Medicine; Chan Zuckerberg Biohub, San Francisco, CA

## Abstract

**IMPORTANCE:** An unhealthy diet is a key risk factor for chronic diseases including obesity, diabetes, and heart disease. Limited access to healthy food options may contribute to unhealthy diets. However, previous studies of food environment have led to mixed results, potentially due to methodological limitations of small sample size, single location, and non-uniform design across studies.

**OBJECTIVE:** To quantify the independent impact of fast food and grocery access, income and education on food consumption and weight status.

**DESIGN, SETTING AND PARTICIPANTS:** Retrospective cohort study of 1,164,926 participants across 9,822 U.S. zip codes logging 2.3 billion consumed foods. Participants were users of the MyFitnessPal smartphone application and used the app to monitor their caloric intake for an average of 197 days each (min 10, max 1,825 days, STD=242).

**MAIN OUTCOMES AND MEASURES:** The primary outcomes were relative change in consumption of fresh fruits and vegetables, fast food, and soda, as well as relative change in likelihood of overweight/obese body mass index (BMI), based on food consumption logs. Food access measures for each zip code were computed from USDA Food Access Research Atlas and Yelp.com, and demographic, income and education measures were based on Census data. Genetic Matching-based approaches were used to create matched pairs of zip codes.

**RESULTS:** Access to grocery stores, non-fast food restaurants, income, and education were independently associated with healthier food consumption and lower prevalence of overweight/obese BMI levels. Substantial differences were observed between predominantly Black, Hispanic, and White zip codes. For instance, within predominantly Black zip codes we found that high income was associated with a *decrease* in healthful food consumption patterns across fresh fruits and vegetables and fast food. Further, high grocery access had a significantly larger association with increased fruit and vegetable consumption in predominantly Hispanic (7.4% increase) and Black (10.2% increase) zip codes in contrast to predominantly White zip codes (1.7% increase).

**CONCLUSIONS AND RELEVANCE:** Policy targeted at improving access to grocery stores, access to non-fast food restaurants, income and education may significantly increase healthy eating, but interventions may need to be adapted to specific subpopulations for optimal effectiveness.

**Note:** We will release all data aggregated at a zipcode level in order to enable validation, follow-up research, and use by policy makers.

**Key Points:** *Question:* How does food consumption and weight status vary with food access, income and education in the United States?

*Findings:* In this country-wide observational study of 1,164,926 participants and 2.3 billion food entries, higher access to grocery stores, lower access to fast food, higher income and education were independently associated with higher consumption of fresh fruits and vegetables, lower consumption of fast food and soda, and lower likelihood of being overweight/obese, but these associations varied significantly across Black, Hispanic, and White subpopulations.

*Meaning:* Policy targeted at improving food access, income and education may increase healthy eating, but interventions may need to be targeted to specific subpopulations for optimal effectiveness.

## 1. Introduction

Unhealthy diet is the leading risk factor for disability and mortality globally^5^. Emerging evidence suggests that the built and food environment, behavioral, and socioeconomic cues and triggers significantly affect diet^6^. Prior studies of the food environment and diet have led to mixed results^6–22^, and very few used nationally representative samples. These mixed results are potentially due to methodological limitations of small sample size, differences in geographic contexts, study population, and non-uniform measurements of both the food environment and diet across studies. Therefore, research with larger sample size and using improved and consistent methods and measurements is needed^8, 23, 24^. With ever increasing smartphone ownership in the U.S.^25^ and the availability of immense geospatial data, there are now unprecedented opportunities to combine various data on individual diets, population characteristics (gender, race and ethnicity), socioeconomic status (income and education), as well as food environment at large scale. Interrogation of these rich data resources to examine geographical and other forms of heterogeneity in the effect of food environments on health could lead to the development and implementation of cost-effective interventions^26^. Here, we leverage large-scale smartphone-based food journals and combine several Internet data sources to quantify the independent impact of food (grocery and fast food) access, income and education on food consumption and weight status of 1,164,926 subjects across 9,822 U.S. zip codes. This study constitutes the largest nationwide study examining the impact of the food environment on diet to date, with 300 times more participants and 4 times more person years of tracking than the Framingham Heart Study^1^.

## 2 Methods

### 2.1 Study Design and Population

We conducted a United States countrywide cross-sectional study of participants’ self-reported food intake and body-mass index (BMI) in relation to demographic (education, ethnicity), socioeconomic (income), and food environment factors (grocery store and fast food access) captured on zip code level.

Overall, this retrospective cohort study analyzed 2.3 billion food intake logs from U.S. smartphone participants over seven years across 9,822 zip codes (U.S. has total of 41,685 zip codes). Participants were participants of the MyFitnessPal app, a free application for tracking caloric intake. We analyzed anonymized, retrospective data collected during a 7-year observation period between 2010 and 2016 that were aggregated to the zip code level. Supplementary Table 1 includes basic statistics on study population demographics and weight status (Body Mass Index; BMI). Data handling and analysis was conducted in accordance with the guidelines of the Stanford University Institutional Review Board.

### 2.2 Study Data

We compute outcome measures of food consumption and weight status from billion food intake logs by 1,164,926 U.S. participants of the MyFitnessPal (MFP) smartphone application to quantify food consumption across 9,822 zip codes. During the observation period from January 1, 2010 to November 15, 2016, the average participant logged 9.30 entries into their digital food journal per day. The average participant used the app for 197 days. All participants in this sample used the app for at least 10 days.

We obtained data on demographic and socioeconomic factors from CensusReporter^27^. Specifically, for each zip code in our data set we obtained median family income, fraction of population with college education (Bachelor’s degree or higher), and fraction of population that is White (not including Hispanic), Black, or Hispanic from the 2010-14 American Community Survey’s census tract estimates^27^.

Grocery store access was defined as the fraction of population that is more than 0.5 miles away from grocery store) following the food desert status definitions from the USDA Food Access Research Atlas^28^. We measured fast food access through the fraction of restaurants that are fast food restaurants within a 40 km (25 miles) radius of the zip code center based on information on Yelp. See Supplementary information for details and validation of these objective food environment measures.

We will release all data aggregated at a zipcode level in order to enable validation, follow-up research, and use by policy makers.

### 2.3 Reproducing state-of-the-art measures using population-scale digital food logs

To investigate the applicability of population-scale digital food logs to study the impact of food environment, income and education on food consumption, we measured the correlation between our smartphone app-based measures and state-of-the-art measures of food consumption including the Behavioral Risk Factor Surveillance System (BRFSS), based on representative surveys, and the Nielsen Homescan data, which is a commercial food purchase database (Figure 3).

**Figure 1:**
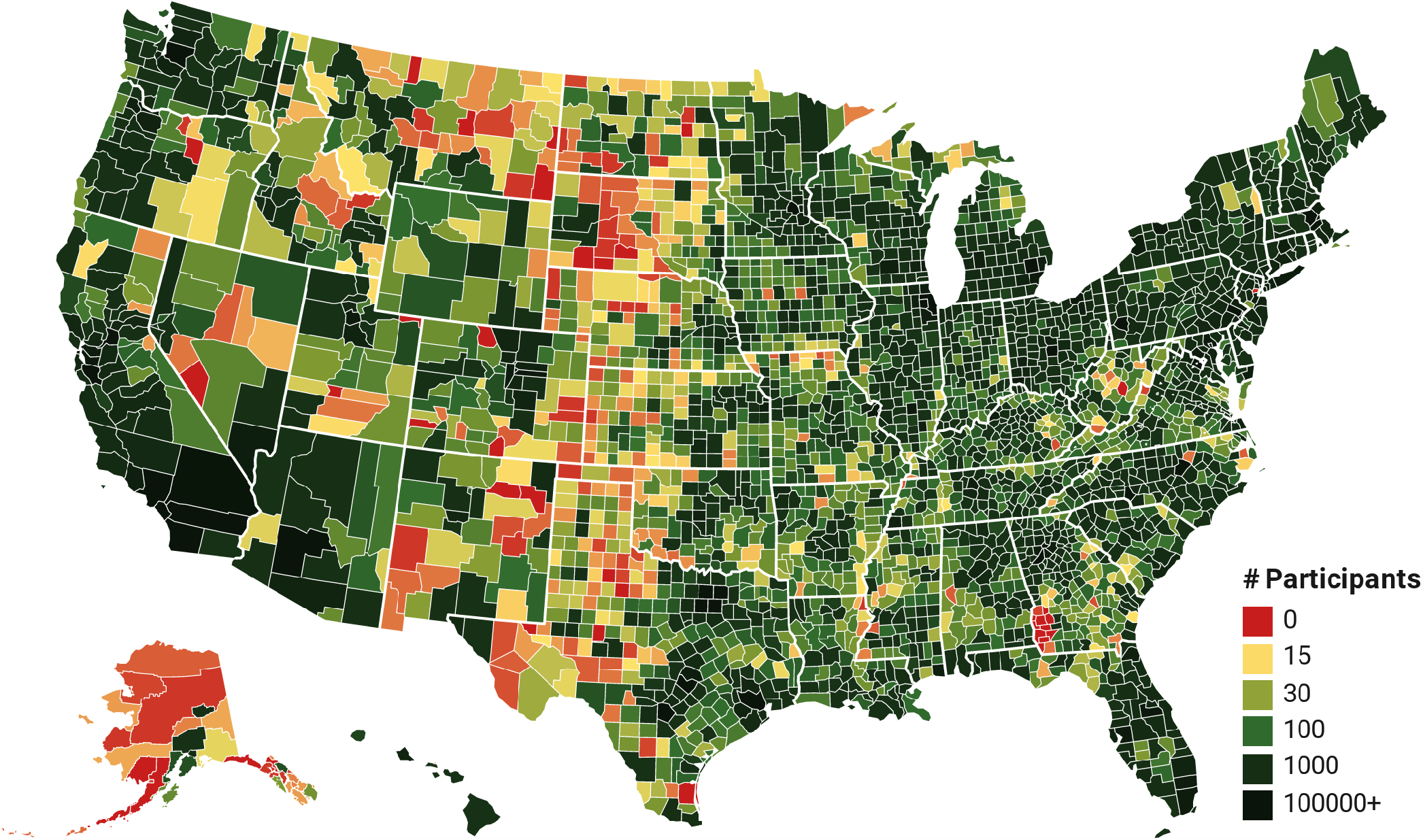
Number of participants in our study across U.S. counties. A choropleth showing the number of participants in each U.S. county. This country-wide observational study included 1,164,926 participants across 9,822 U.S. zip codes that collectively logged 2.3 billion food entries for an average of 197 days each. Most U.S. counties are represented by at least 30 participants in our dataset. This study constitutes the largest nationwide study examining the impact of food environment on diet to date, with 300 times more participants and 4 times more person years of tracking than the Framingham Heart Study^1^.

**Figure 2:**
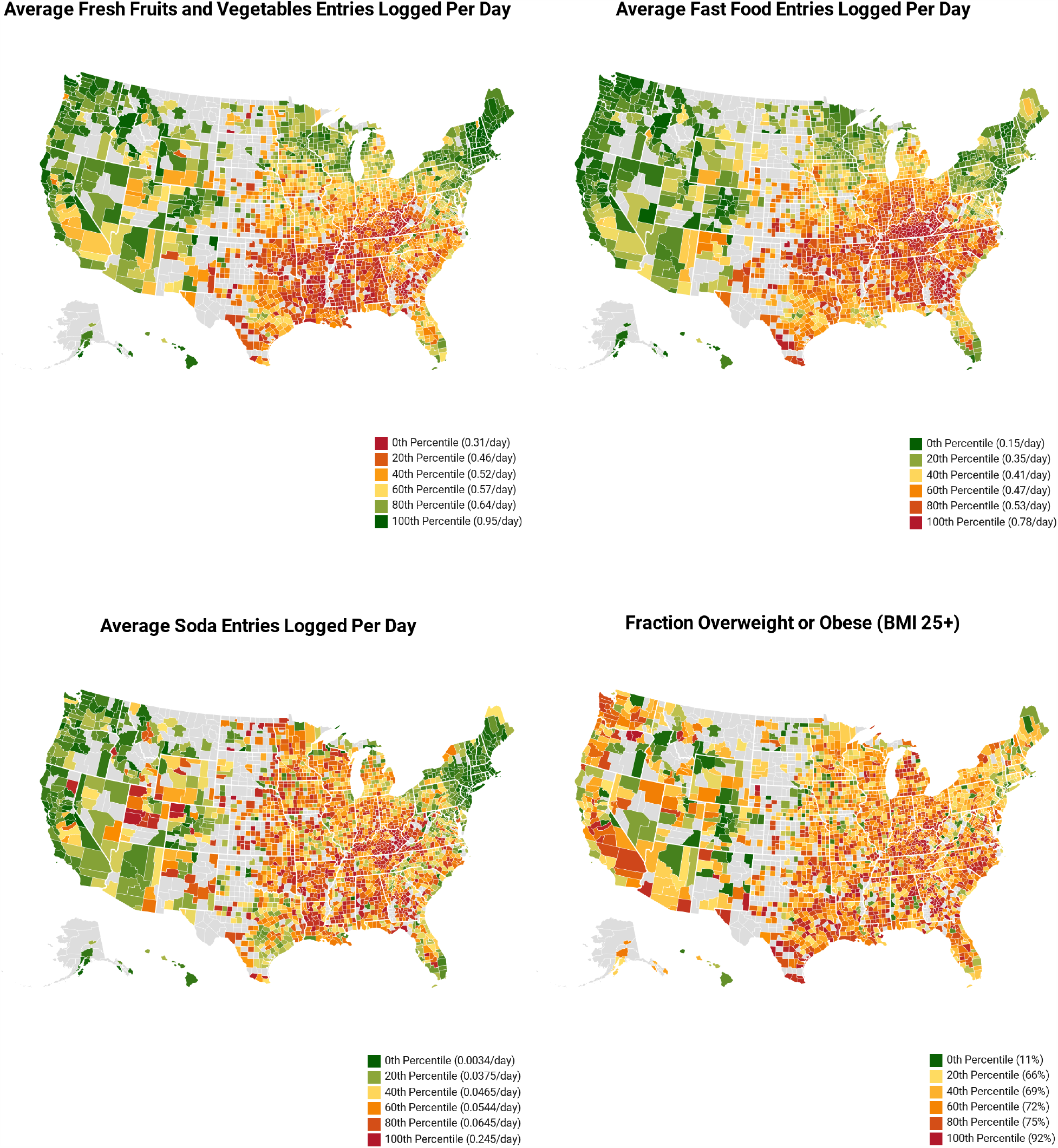
Dietary Consumption and Body Weight across U.S. Counties. A set of choropleths showing the main study outcomes of the number of entries that are classified as fresh fruit and vegetables, fast food, and soda consumption as well as the fraction of overweight/obese participants across the USA by counties with more than 30 participants. We observe that food consumption healthfulness varies significantly across counties in the United States.

**Figure 3:**
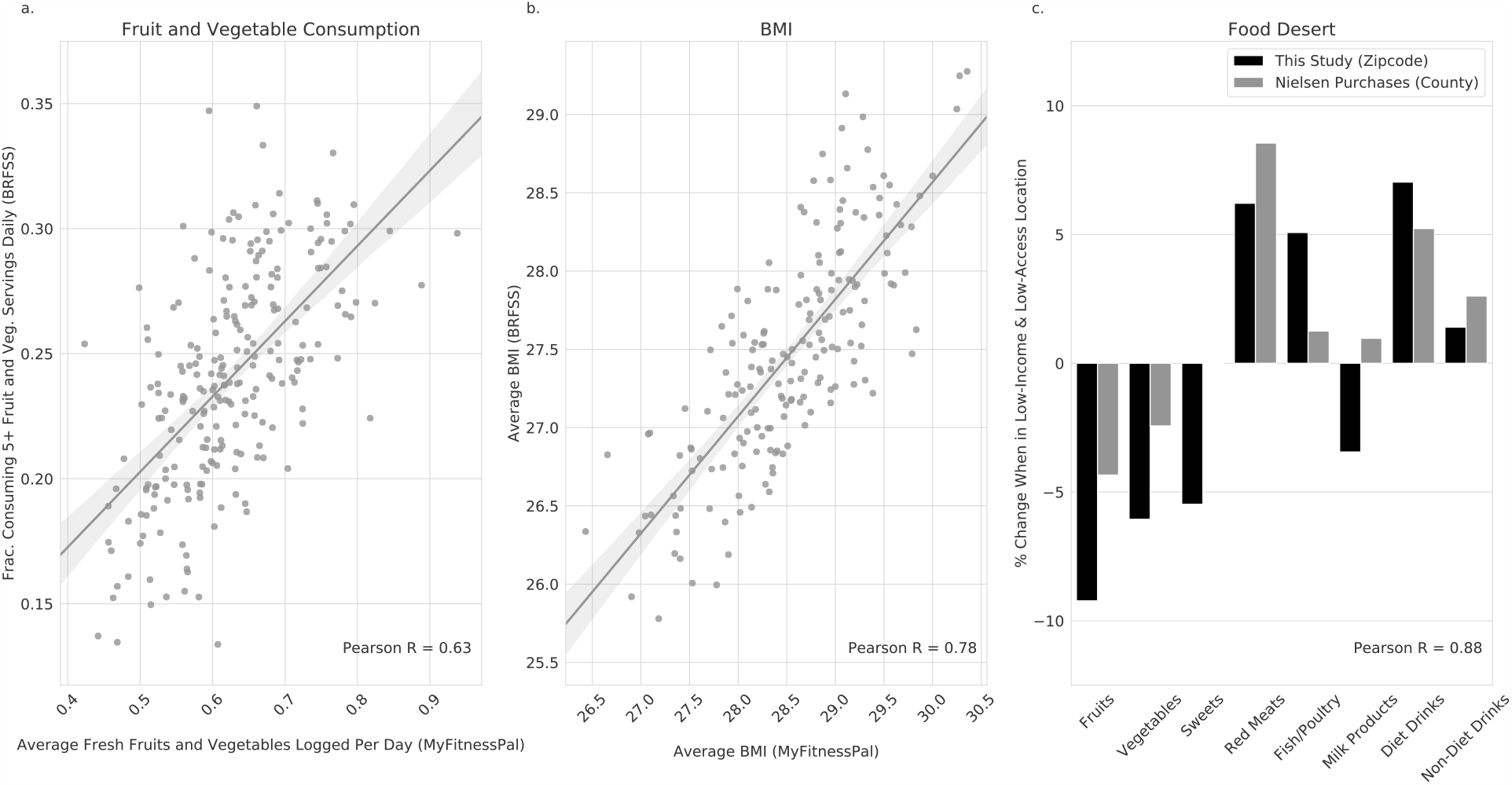
This studies’ smartphone-based food logs correlate with previous, representative survey measures and purchase data. **a**, Fraction of fresh fruits and vegetables logged is correlated with BRFSS survey data^2^ (R=0.63, *p* < 10^−5^; Methods). **b**, Body Mass Index of smartphone cohort is correlated with BRFSS survey data^3^ (R=0.78, *p* < 10^−5^; Methods). **c**, Digital food logs replicate previous findings of relative consumption differences in low-income, low-access food deserts based on Nielsen purchase data^4^ (R=0.88, *p* < 0.01; Methods). These results demonstrate that smartphone-based food logs are highly correlated with existing, gold-standard survey measures and purchase data.

Comparing our data to BRFSS on county level, we found strong correlations between the amount of fresh fruits and vegetables (F&V) consumed (Figure 3a, R=0.63, *p* < 10^−5^) and body mass index (Figure 3b, R=0.78, *p* < 10^−5^). Comparing to USDA purchase data from the Nielsen Homescan Panel Survey we found that our app-based food logs were very highly correlated with previously published results (Figure 3c, R=0.88, *p* < 0.01) and that the absolute differences between food deserts and non-food deserts were stronger in the MFP data compared to Nielsen purchase data. See Supplementary information for more details. These results demonstrate convergent validity and suggest that population-scale digital food logs can reproduce the basic dynamics of traditional, state-of-the-art measures, and they can do so at massive scale and comparatively low cost.

### 2.4 Statistical Analysis

In this large-scale observational study, we used a matching-based approach^29, 30^ to disentangle contributions of income, education, grocery access, and fast food access on food consumption. To estimate the impact of each of these factors, we divide all available zip codes into treatment and control groups based on a median split; that is, we estimate the difference in outcomes between matched above-median and below-median zip codes. We create matched pairs of zip codes by selecting a zip code in the control group that is closely matched to the zip code in the treatment group across all factors, except the treatment factor of interest. Since we repeat this matching process for each zip code in the treatment group, this approach estimates the Average Treatment Effect on the Treated (ATT). Through this process, we attempt to eliminate variation of plausible influences and to isolate the effect of interest. We repeat this process for each treatment of interest; for example for the results presented in Figure 4, we performed four matchings, one for each of income, education, grocery access and fast food access. For the sub- population experiments (Figure 5), we repeated the same method on the subset of the zip codes in which the majority of inhabitants were of a particular racial group. See Supplementary information for details on the matching approach and detailed statistics that demonstrate that treatment and control groups were well-balanced on observed covariates after matching.

**Figure 4:**
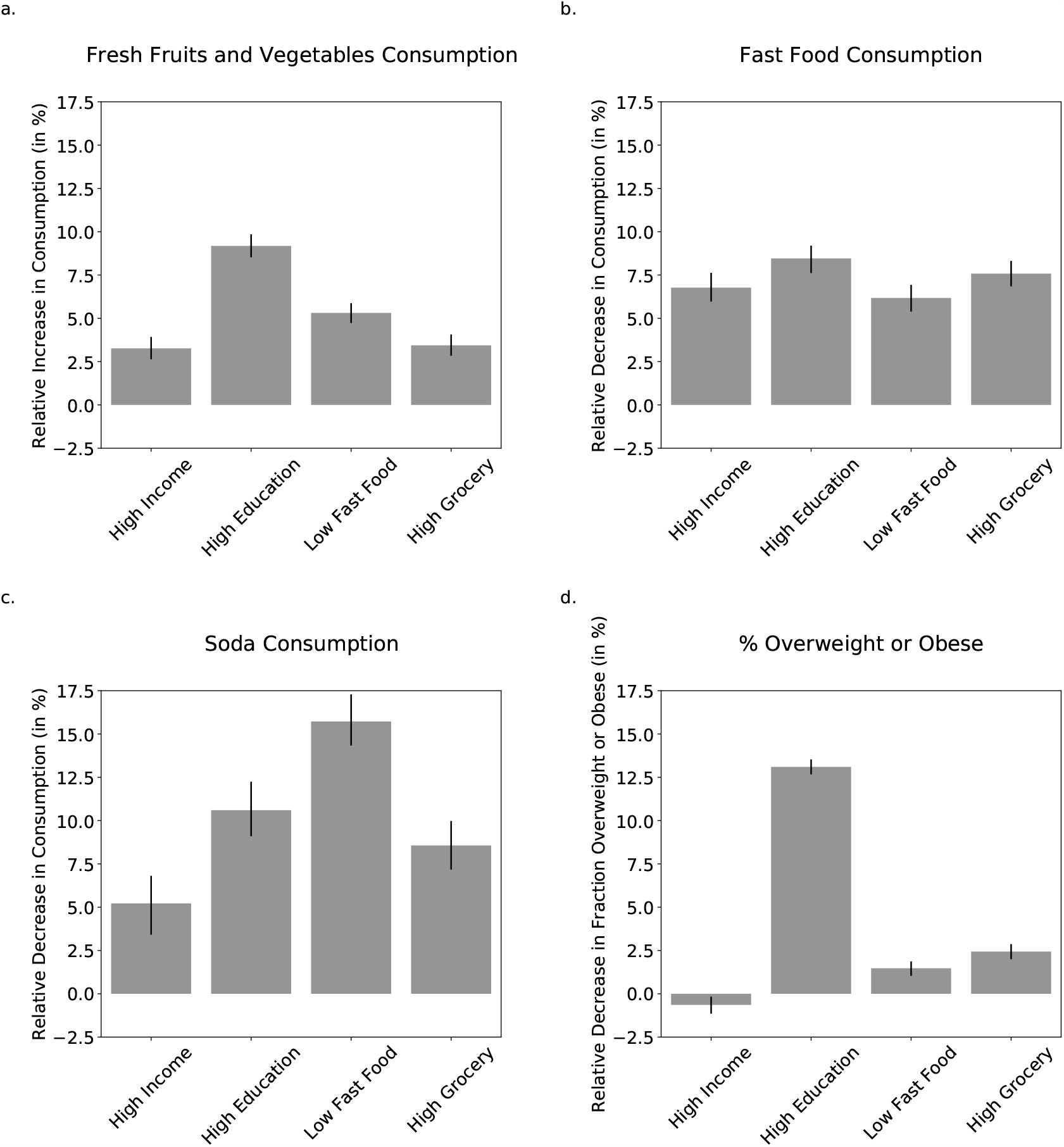
The impact of income, education, grocery store and fast food access on food consumption and weight status. Independent contributions of high income (median family income higher than or equal to $70,241), higher education (fraction of population with college education 29.8% or higher), high grocery access (fraction of population that is closer than 0.5 miles from nearest grocery store is greater than or equal to than 20.3%), and low fast food access (less than or equal to 5.0% of all businesses are fast-food chains) on relative change in consumption of **a**, fresh fruits and vegetables, **b**, fast food, **c**, soda, and **d**, relative change in percent overweight or obese (BMI 25+). Cut points correspond to median values. Estimates are based on matching experiments controlling for all but the one treatment variable in focus (Methods). Error bars correspond to bootstrapped 95% confidence intervals (Supplementary Methods). While the most impactful factors vary across outcomes, only higher education was associated with a sizeable decrease of 13.1% in overweight and obese weight status.

**Figure 5:**
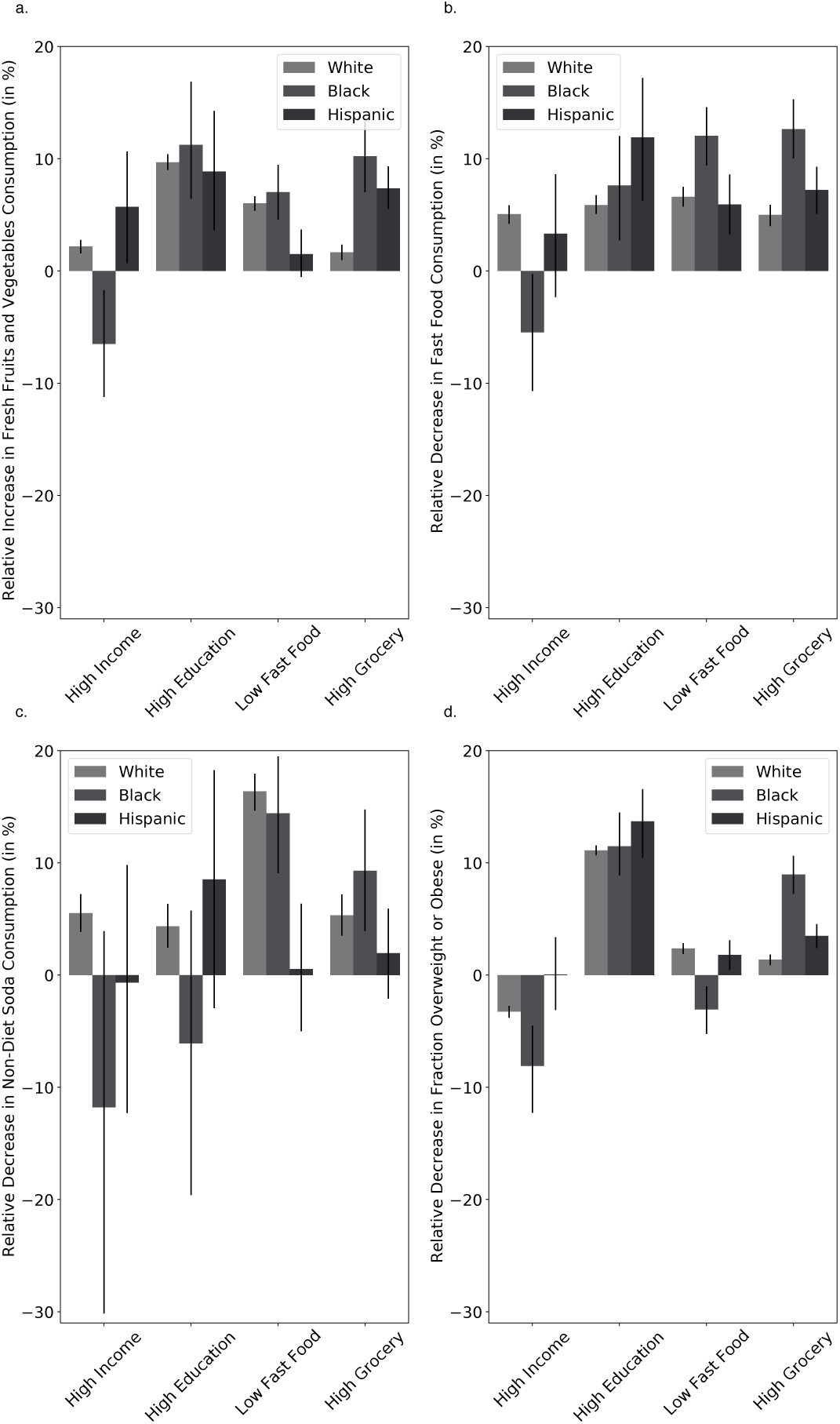
Effects on food consumption and weight status disaggregated by predominantly Black, Hispanic, and non-Hispanic White zip codes (*i*.*e*., 50% or more). Independent contributions of high income (median family income higher than or equal to $70,241), higher education (fraction of population with college education 29.8% or higher), high grocery access (fraction of population that is closer than 0.5 miles from nearest grocery store is greater than or equal to than 20.3%), and low fast food access (less than or equal to 5.0% of all businesses are fast-food chains) on relative change in consumption of **a**, fresh fruits and vegetables, **b**, fast food, **c**, soda, and **d**, relative change in percent overweight or obese (BMI > 25). Cut points correspond to median values. Estimates are based on matching experiments controlling for all but one treatment variable (Methods). Error bars correspond to bootstrapped 95% confidence intervals (Methods). We observe significant differences across Black, Hispanic, and non-Hispanic White zip codes in terms how food consumption is affected by factors of income, education, fast food and grocery access.

We tested discriminant validity of our statistical approach by measuring the effect of null- treatments that should not have any impact on food consumption. We chose examples of null- treatments by selecting variables that had little correlation with study independent variables (income, education, grocery access, fast food access) and were plausibly unrelated to food consumption. This selection process lead to use of the fraction of countertop installers, electronics stores, and waterproofing services nearby as measured through Yelp. Applying our analysis pipeline to these null-treatments, we found that all of these null-treatments had zero effect on food consumption. This demonstrated that our statistical analysis approach did not produce measurements that it was not supposed to measure; that is, discriminant validity (Figure 7).

**Figure 7:**
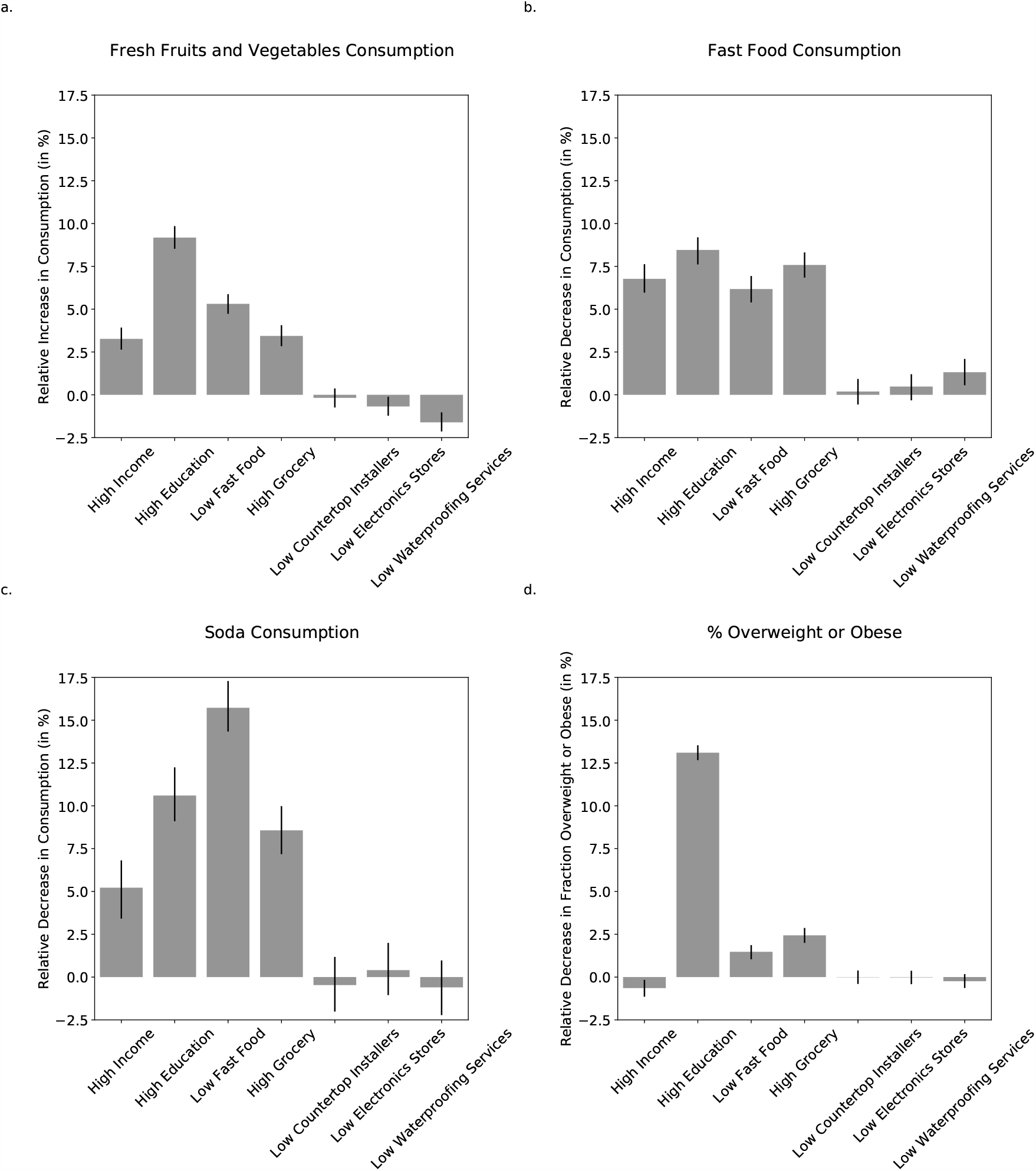
Demonstration of discriminant validity of statistical approach. We measured the effect of null-treatments that should not have any impact on food consumption. We chose examples of null-treatments by selecting variables that had little correlation with study independent variables (income, education, grocery access, fast food access) and were plausibly unrelated to food consumption. This selection process lead to use of the fraction of countertop installers, electronics stores, and waterproofing services nearby as measured through Yelp. Applying our analysis pipeline to these null-treatments, we found that all of these effect estimates were close to zero. This demonstrated that our statistical analysis approach did not produce measurements that it was not supposed to measure; that is, discriminant validity.

## 3. Results

Across all 9,822 U.S. codes, we found that high income, high education, high grocery access, and low fast food access were independently associated with higher consumption of fresh fruits and vegetables (F&V), lower consumption of fast food and soda, and lower prevalence of overweight or obese BMI levels (Figure 4).^1^ Specifically, in zip codes of above median grocery access participants logged 3.4% more F&V, 7.6% less fast food, 8.6% less soda and were 2.4% less likely to be overweight or obese (all *P* < 0.001). In zip codes of below median fast food access participants logged 5.3% more F&V, 6.2% less fast food, 15.7% less soda and were 1.5% less likely to be overweight or obese (all *P* < 0.001). In zip codes of above median education participants logged 9.2% more F&V, 8.5% less fast food, 10.6% less soda and were 13.1% less likely to be overweight or obese (all *P* < 0.001). Finally, in zip codes of above median household income (referred to as *higher income* below), participants logged 3.3% more F&V, 6.8% less fast food, 5.2% less soda (all *P* < 0.001), but had a 0.6% higher likelihood of being overweight or obese (*P* = 0.006). However, above median household income was associated with a 0.34% decrease in BMI (*P* < 0.001). Note that the reported effect size are based on comparing above and below median zip codes for any given factor. We found a general pattern of consistent but increased effect sizes when comparing top versus bottom quartiles (Supplementary Information Figure 6), suggesting a dose-response relationships across all considered variables (with the exception of the association between low fast food access and likelihood of overweight or obese BMI levels). We found that zip codes with high education levels compared to low education levels had the largest relative increases in F&V, fast food, and overweight or obese BMI levels. However, in terms of its impact on soda consumption, we found low fast food access to be associated with the largest relative decrease in soda consumption. In particular, high education zip codes had the largest relative decrease in reported soda consumption (15.7%).

**Figure 6:**
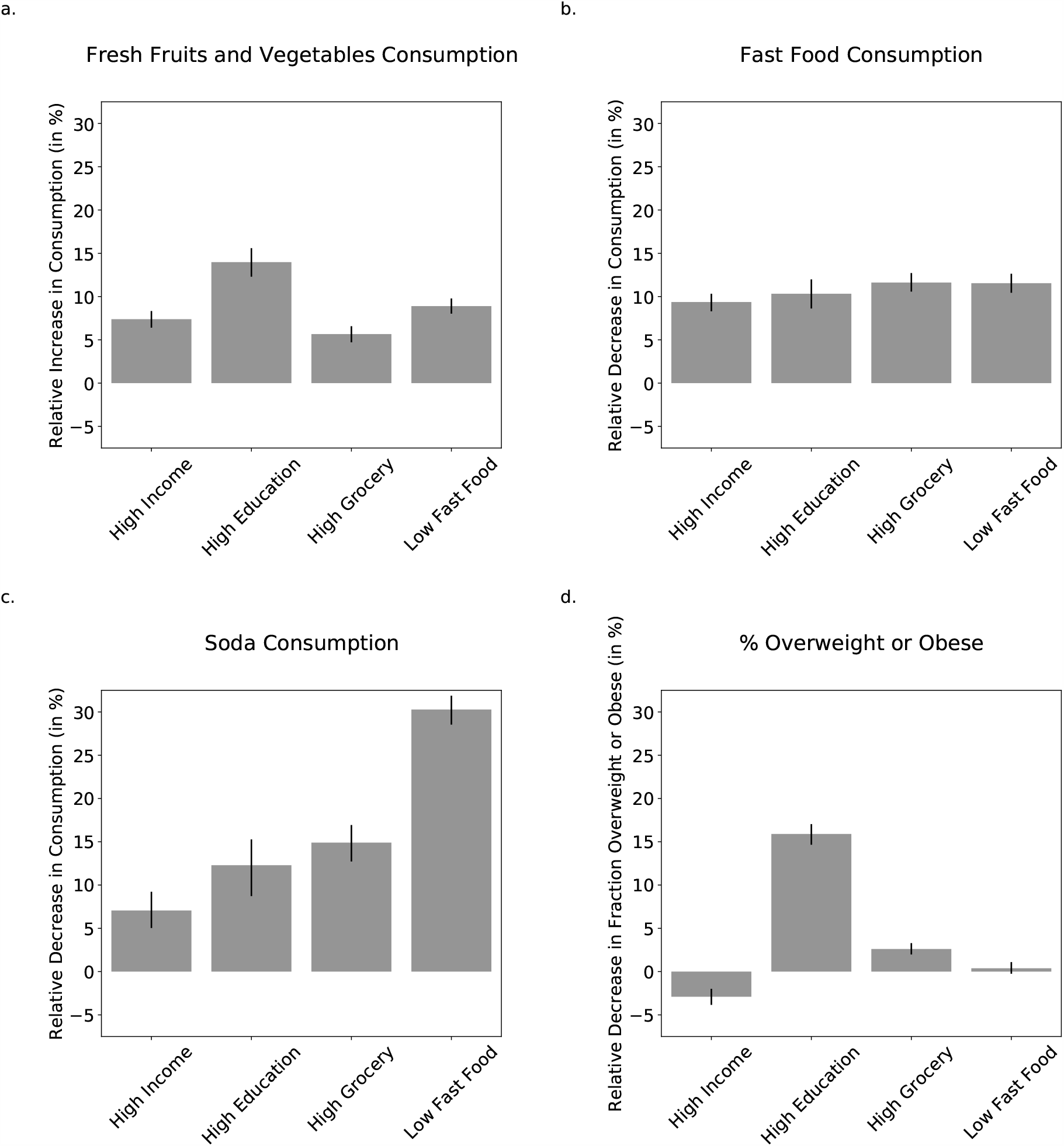
Matching experiments using quartile instead of median split. Note the consistent but increased effect sizes compared to Figure 4.

We separately repeated our data analyses within zip codes that were predominantly Black (3.7%), Hispanic (5.6%) and non-Hispanic White (78.4%) (Figure 5). Results within predominantly non-Hispanic White zip codes closely matched results within the overall population, since most zip codes in this study were predominantly White (78.4%), not unlike to the overall U.S. population (61.3%)^31^. However, restricting our analyses to predominantly Black and Hispanic zip codes led to remarkably different findings. Specifically, within predominantly Black zip codes we found an impact of higher income in the inverse direction of the population average and towards low healthful food consumption, across across four out of four outcome variables, resulting in decreased F&V consumption (6.5%), increased fast food consumption (7.0%), and increased likelihood of overweight or obese BMI levels (8.1%). Higher income was also associated with increased soda consumption (11.8%) but was not statistically significant (*P* = 0.081). On the other hand, low fast food access and high education access were generally associated with increased diet health, with low fast food access correlating with the highest decrease in soda consumption (14.4%) and high education with the highest increase in fresh fruit and vegetable consumption (11.2%), although decreased fast food access was harmful to one of the outcome variables. Specifically, decreased fast food access was associated with a slight increase in fraction overweight or obese (3.1%). The only variable that had a positive effect on the health of all outcome variables was increased grocery access, which was associated with increased F&V consumption (10.2%), decreased fast food consumption (12.6%), decreased soda consumption (9.3%), and decreased likelihood of overweight or obese BMI levels (9.0%).

In contrast, within predominantly Hispanic zip codes we found a significant effect of higher, above-median, income on higher F&V consumption (5.7%), but not across the remaining three outcome variables. Higher education zip codes had the most positive association with diet health across all variables, with the exception of soda consumption for which none of the factors had a significant impact. Specifically, higher education was associated with increased F&V consumption (8.9%), decreased fast food consumption (11.9%), and decreased likelihood of overweight or obese BMI levels (13.7%). Higher grocery access and decreased fast food access had similar effects as on the overall population for some outcome variables (i.e. similar associations with likelihood of overweight or obese BMI levels and fast food consumption). However, in some cases the magnitude of impact was higher (i.e. grocery access increased Hispanic F&V consumption by 7.4% which more than twice the increase of the overall population) and in others, unlike the overall population, there was no significant association (i.e. no significant relationship between fast food/grocery access on soda consumption, or between fast food access and F&V consumption).

Few factors led to consistent improvements across all three subpopulations. Across all three groups, F&V consumption was strongly increased by high grocery access and education. Fast food consumption was reduced by all potential intervention targets besides increased income. Soda consumption was most reduced by decreased fast food access for Black and White-majority zip codes, but was not impacted by any of the intervention targets within Hispanic zip codes. Lastly, overweight or obese BMI levels were, by far, most strongly reduced across all groups by increased education levels.

## 4. Discussion

While many of our results were consistent with previous studies^32–34^, importantly, we found high education zip codes had the largest relative decrease in the likelihood of overweight and obese BMI levels of 13.1%. This suggests that having more education is the most effective at reducing overweight and obesity for all races. Our calculation indicated that a 13.1% decrease in overweight and obesity by implementing effective education programs and policies could potentially lead to $53.6 billion in annual health care cost savings (Supplementary Methods), and could support 83.8% of the education budget for the entire U.S.^35–39^.

Similarly, having higher grocery access, and lower fast food access could potentially lead to $9.9 billions and $6.1 billions of annual health care cost savings, respectively (Supplementary Methods). While it is challenging to close the education and income gaps, establishing more grocery stores and limiting fast food restaurants together could potentially save $16 billion. These changes may work in synergistic ways that may lead to even lower obesity prevalence and obesity-related cost savings. This synergy could be multiplied when combining effective education programs that could potentially lower obesity prevalence further by increasing individuals’ SES (e.g., income and education)^40, 41^, health literacy and behaviors^40–44^, as well as sense of control and empowerment^45^.

When we restricted our analyses to predominantly Black and Hispanic zip codes, we found the independent impacts of food access, income and education on food consumption and weight status varied significantly across Black, Hispanic and White populations. These findings suggest that tailored intervention strategies are needed based on neighborhood population distributions, assets and contexts.

Our results showed that the impact of having higher grocery store access significantly increased fresh fruit and vegetable consumption and decreased fast food consumption for both Blacks and Hispanics much more than for Whites. Although previous literature has shown null effects of grocery access^46, 47^, and argued against existing government policies that tried to increase grocery access in certain areas^48, 49^, these studies have focused on the general population, which is White- skewed. Therefore, policies and strategies in increasing grocery store access and decreasing fast food access could potentially be the most effective approaches in changing dietary habits among people of color.

Within predominantly Black zip codes, unlike other races, the effect of higher education was low compared to grocery and fast food access. Having higher income was associated with decreased fruits and vegetable consumption, increased fast food and soda consumption, and increased likelihood of overweight and obesity. This could be explained by the diminishing return hypothesis, which suggests that Blacks receive fewer protective health benefits from increases in SES than Whites^50, 51^. A combination of factors, including neighborhood economic disadvantage^52, 53^, racial discrimination^54, 55^, and stress associated with educational attainment and mobility^56^, may prevent higher SES Blacks from achieving their fullest health potential relative to Whites^57^.

Within predominantly Hispanic zip codes, higher income was not associated with reduced likelihood of overweight or obese BMI levels, but higher education significantly reduced that likelihood by 13.7%. High grocery access was the only factor associated with higher consumption of fresh fruits and vegetables, and high grocery and low fast food access had similar effects in reducing fast food and soda consumption and lowering overweight and obese BMI levels. The relationship between higher income and BMI could be partially explained by the “Hispanic health paradox” and “Hispanic health advantage”^58–63^. Additionally, through acculturation or adopting American culture, Hispanic immigrants may engage in less healthy behaviors, which in turn put themselves at higher risk for chronic diseases^58–60, 64–68^.

Due to the cross-sectional nature of the study, we were not able to make any causal inferences between SES and food environment variables and dietary behavior and BMI. However, we used a matching-based approach to mimic a quasi-experimental design to disentangle the individual impact of income, education and food access on participants food consumption. Our analysis did not include other demographic variables such as gender and age, but we jointly considered the impacts of income, education and food environment access on participants food consumption with consistent measures across the U.S., whereas previously published studies examined one or a few at a time. We used individuals food loggings to estimate their consumption. Food loggings may not capture what individuals actually ate. However, we conducted rigorous validations through comparisons with high quality and highly representative datasets which demonstrated high correlations to gold-standard approaches (Figure 3). Majority of the food environment studies used screeners, food frequency questionnaires or 24-hour recalls for dietary assessment, and very few used diaries^8^. Our participants logged their food intakes for an average of 197 days each. We also harnessed other large datasets such as Yelp to examine participants food environments. Considering both the strengths and limitations of this study, more research is needed especially based on longitudinal study design and detailed individual level data to allow causal inference and precise interpretation of the results.

## 5. Conclusion

We analyzed 2.3 billion food intake logs and BMIs from 1.2 million MyFitnessPal smartphone app participants over seven years across 9,822 zip codes in relation to education, race/ethnicity, income, and food environment access. Our analyses indicated that higher access to grocery stores, lower access to fast food, higher income and education were independently associated with higher consumption of fresh fruits and vegetables, lower consumption of fast food and soda, and lower likelihood of being overweight or obese. Policy targeted at improving food access, income and education may increase healthy eating. Potential interventions may need to be targeted to specific subpopulations for optimal effectiveness.

## Data Availability

Note We will release all data aggregated at a zipcode level in order to enable validation, follow-up research, and use by policy makers.

## ARTICLE INFORMATION

### Author Affiliations

Allen School of Computer Science & Engineering, University of Washington (Tim Althoff); Department of Computer Science, Stanford University (Hamed Nilforoshan); Stanford Prevention Research Center, School of Medicine, Stanford University (Jenna Hua). Department of Computer Science, Stanford University (Jure Leskovec);

### Author Contributions

Study concept and design: Althoff and Leskovec. Statistical analysis: Althoff and Nilforoshan. Interpretation of data: All authors. Drafting of the manuscript: All authors. Critical revision of the manuscript for important intellectual content: All authors.

## Funding/Support

T.A. and J.L. were supported by a National Institutes of Health (NIH) grant (U54 EB020405, Mobilize Center, NIH Big Data to Knowledge Center of Excellence). T.A. was supported by the SAP Stanford Graduate Fellowship, NSF grant IIS-1901386, and Bill Melinda Gates Foundation (INV-004841). H.N was supported by NSF REU #1659585 at the Stanford Center for the Study of Language and Information (CSLI). J.H is supported by Postdoctoral Fellowship in Cardiovascular Disease Prevention (T32) funded by the National Heart, Lung, and Blood Institute (NHLBI) at the National Institutes of Health (NIH). J.L. was supported by DARPA under Nos. FA865018C7880 (ASED), N660011924033 (MCS); ARO under Nos. W911NF-16-1-0342 (MURI), W911NF-16-1-0171 (DURIP); NSF under Nos. OAC-1835598 (CINES), OAC-1934578 (HDR), CCF-1918940 (Expeditions); Stanford Data Science Initiative, Wu Tsai Neurosciences Institute, Chan Zuckerberg Biohub, Amazon, Boeing, Chase, Docomo, Hitachi, Huawei, JD.com, NVIDIA, Dell.

### Roles of Funder/Sponsor

The funding sources had no role in the design and conduct of the study; collection, management, analysis, and interpretation of the data; preparation, review, or approval of the manuscript; and decision to submit the manuscript for publication.

### Disclaimer

The content is solely the responsibility of the authors and does not necessarily represent the official views of the NIH or sponsors.

### Additional Contributions

We thank MyFitnessPal for donating the data for independent research and Mitchell Gordon for helping with data preparation.

## Supplementary Information

### Supplementary Methods

#### Details on food environment measures

We obtained data on grocery store access (fraction of population that is more than 0.5 miles away from grocery store) and food desert status from the USDA Food Access Research Atlas^28^. A census tract is considered a food desert by the USDA if it is both low-income (defined by Department of Treasurys New Markets Tax Credit program) and low-access, meaning at least 500 people or 30 percent of residents live more than 1 mile from a supermarket in urban areas (10 miles in rural areas)^4^. We aggregated these data from a census tract level to a zip code level using USPS Crosswalk data, which provides a list of all census tracts which overlap with a single zip-code^75^. We related these data on census tract level to the zip code level by taking the weighted average of each census tract food environment measure (both grocery store access and food desert status), weighted by the number of people in the tract^75^. For instance, if zip code A overlapped with Census Tract A (2500 people, food desert) and Census Tract B (7500 people, not a food desert), the food desert measure of zip code A would be estimated as 25%. We defined the binary threshold for food desert, used in Figure 3, as 50% or higher.

We measured fast food access through the fraction of restaurants that are fast food restaurants within a 40 km (25 miles) radius of the zip code center. Data on local restaurants and businesses were obtained through the Yelp API^76^. For each zip code, we consider up to 1000 restaurant businesses that are nearest to the zip code center up to a distance of 40km (67.8% of zip code queries resulted in 1000 restaurant businesses within 40km; Yelp API results are restricted to 1000 results). Zip codes with more than 1000 restaurant businesses within 40km are likely more dense urban environments and we still capture the nearest restaurant businesses to the zip code center that arguably capture the local food environment. We further used Yelp-based environment variables that we expected *not* to influence food consumption, such as the availability of waterproofing services, countertop installers, or electronic stores, as null experiments to demonstrate discriminant validity of our statistical analysis pipeline (see Supplementary Figure 7).

#### Details on outcome measure (food consumption and weight status)

We used 2.3 billion food intake logs by 1,164,926 U.S. participants of the MyFitnessPal (MFP) smartphone application to quantify food consumption across 9,822 zip codes. During the observation period from January 1, 2010 to November 15, 2016, the average participant logged 9.30 entries into their digital food journal per day. The average participant used the app for 197 days. All participants in this sample used the app for at least 10 days. Each entry contains a separate food component (*e*.*g*., banana, yogurt, hamburger, …). We classified all entries into three categories: fresh fruits and vegetables (F&V; through a proprietary classifier by MFP), fast food (if the description contained the name of a fast food chain listed in Supplementary Information Table 6, and sugary (non-diet) soda (if the description contained the name of a soda drink listed in Supplementary Information Table 7 but did not contain “diet”, “lite”, “light”, or “zero”). In all cases, descriptions, as well fast food and soda drink keywords, were normalized by lower-casing and removing punctuation. We then calculated the average number of food entries logged per participant, per day, for each of the F&V, fast food, and soda categories (e.g. average number of F&V logged per participant per day), excluding days in which the participant was inactive (*i*.*e*., consistently did not log anything). Finally, we aggregated these participant-level measures to the zip code level by taking the mean of each category’s measure for all participants in each zip code. We further used body-mass index (BMI) health in each zip code as a weight status outcome. We operationalize BMI health as the fraction of participants in a zip code which are overweight or obese (BMI > 25). BMI was self-reported by participants of the smartphone application (99.92% of participants did report BMI). Table 2 shows basic summary statistics for the outcome measures used in this study. In our statistical analyses, we compared two sets of zip codes that differ in a dimension of interest (*e*.*g*., grocery store access access) as treatment and control group and use the relative change in F&V consumption, fast food consumption, soda consumption, and BMI health of the treated group relative to the control group. To generate confidence intervals, as well as to compute p-values to test for statistical significance of changes in outcome, we use non-parametric bootstrap resampling^77^. Specifically, we follow the method proposed by Austin and Small,^78^ which is to draw bootstrap samples post-matching from the matched pairs in the propensity-score-matched sample after the Genetic Matching stage^79^. We confirmed the validity of this method empirically by also calculating t-tests for each experiment, which gave qualitatively similar results.

**Table 1:** Demographic statistics for our study compared with nationally representative survey data. (*) indicates statistics calculated at the zip code level.

| Source | BMI | Overweight<br>or Obese | Obese | Median Age | Gender | Median<br>Family Income* | College* | Ethnicity* |
| --- | --- | --- | --- | --- | --- | --- | --- | --- |
| Our Study | 28.5 | 67.8% | 32.8% | 36 | 74% Female | \$ 76,563 | 33.7% | 68.3% White |
| Nat.l Avg. | 29.4<sup>69</sup> | 71.6%<sup>70</sup> | 39.8%<sup>70</sup> | 38.2<sup>71</sup> | 50.5% Female<sup>72</sup> | \$ 59,039<sup>73</sup> | 33.4%<sup>74</sup> | 61.3% White<sup>31</sup> |

**Table 2:**
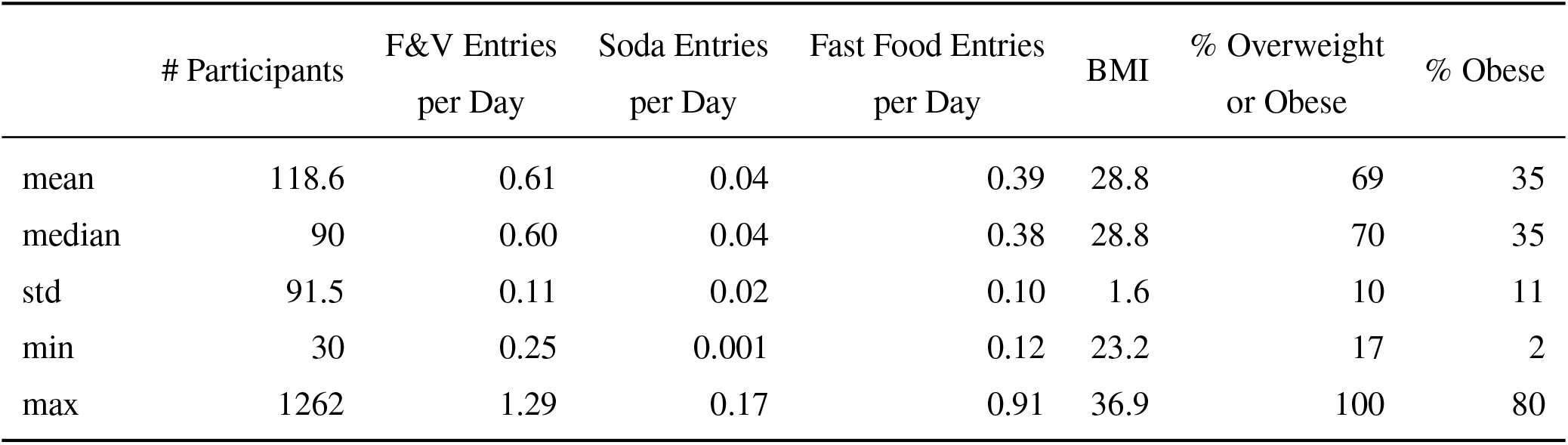
Outcome measures calculated at the zip code level for the **9,822** zip codes in our study, spanning **1,164,926** participants.

**Table 3:**
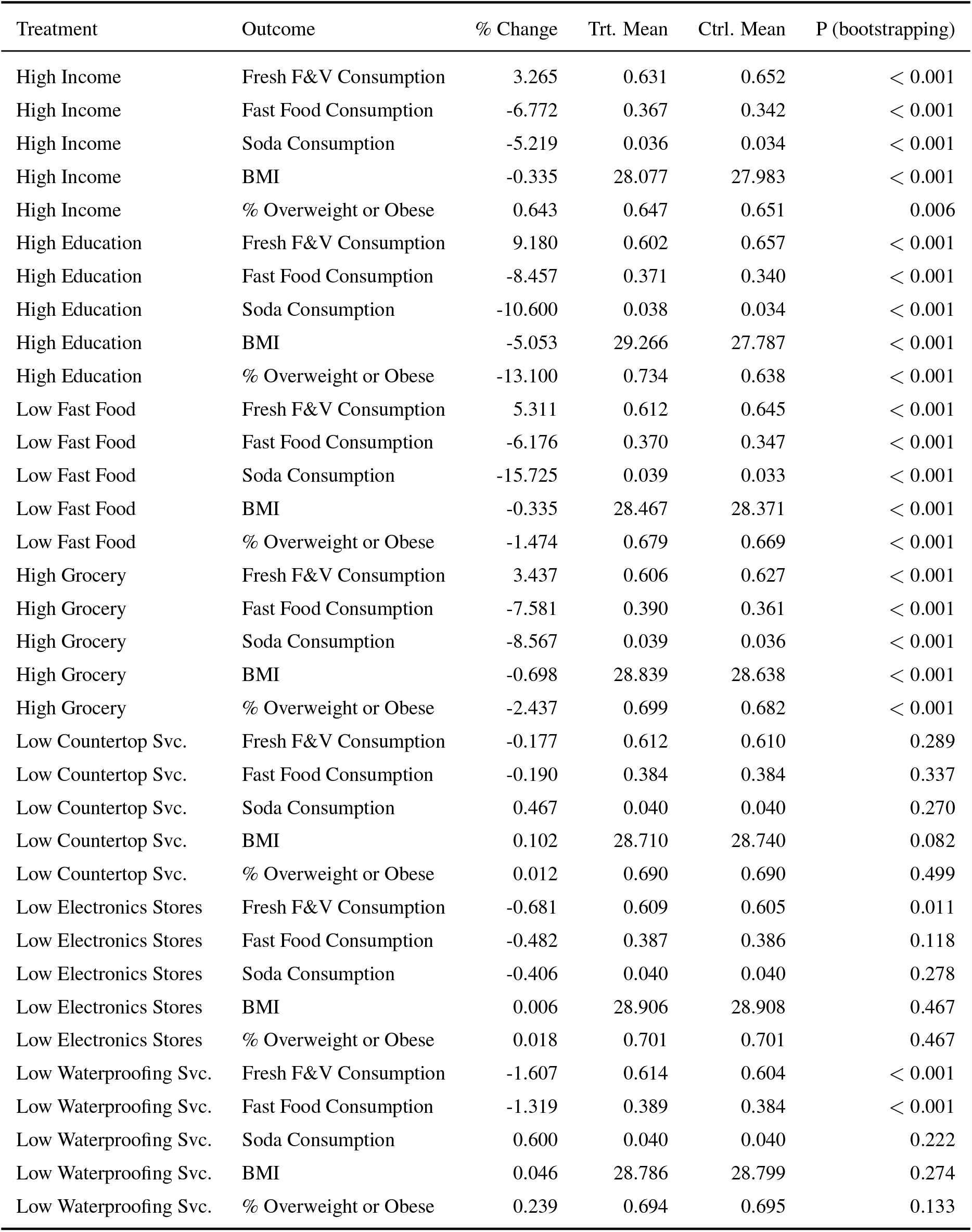
Effect sizes of all top/bottom half matching experiments (Fig. 4)

**Table 4:** Effect sizes of all race-specific top/bottom half matching experiments (Fig. 5).

| Race | Treatment | Outcome | % Change | Trt. Mean | Ctrl. Mean | P-value (bootstrapping) |
| --- | --- | --- | --- | --- | --- | --- |
| Black | High Income | Fresh F&V Consumption | -6.497 | 0.607 | 0.567 | 0.004 |
| Black | High Income | Fast Food Consumption | 5.464 | 0.403 | 0.425 | 0.015 |
| Black | High Income | Soda Consumption | 11.781 | 0.033 | 0.037 | 0.081 |
| Black | High Income | BMI | 3.695 | 29.834 | 30.936 | < 0.001 |
| Black | High Income | % Overweight or Obese | 8.101 | 0.750 | 0.811 | < 0.001 |
| Black | High Education | Fresh F&V Consumption | 11.243 | 0.558 | 0.620 | < 0.001 |
| Black | High Education | Fast Food Consumption | -7.613 | 0.424 | 0.391 | 0.002 |
| Black | High Education | Soda Consumption | 6.088 | 0.033 | 0.035 | 0.150 |
| Black | High Education | BMI | -5.512 | 31.362 | 29.634 | < 0.001 |
| Black | High Education | % Overweight or Obese | -11.481 | 0.829 | 0.734 | < 0.001 |
| Black | Low Fast Food | Fresh F&V Consumption | 7.024 | 0.543 | 0.582 | < 0.001 |
| Black | Low Fast Food | Fast Food Consumption | -12.043 | 0.473 | 0.416 | < 0.001 |
| Black | Low Fast Food | Soda Consumption | -14.421 | 0.040 | 0.034 | < 0.001 |
| Black | Low Fast Food | BMI | 0.506 | 30.776 | 30.931 | 0.153 |
| Black | Low Fast Food | % Overweight or Obese | 3.062 | 0.775 | 0.799 | 0.001 |
| Black | High Grocery | Fresh F&V Consumption | 10.230 | 0.527 | 0.581 | < 0.001 |
| Black | High Grocery | Fast Food Consumption | -12.642 | 0.478 | 0.418 | < 0.001 |
| Black | High Grocery | Soda Consumption | -9.300 | 0.039 | 0.036 | 0.002 |
| Black | High Grocery | BMI | -3.795 | 31.966 | 30.753 | < 0.001 |
| Black | High Grocery | % Overweight or Obese | -8.960 | 0.861 | 0.783 | < 0.001 |
| Hispanic | High Income | Fresh F&V Consumption | 5.706 | 0.556 | 0.588 | 0.012 |
| Hispanic | High Income | Fast Food Consumption | -3.314 | 0.393 | 0.380 | 0.140 |
| Hispanic | High Income | Soda Consumption | 0.663 | 0.036 | 0.036 | 0.429 |
| Hispanic | High Income | BMI | 0.289 | 28.997 | 29.080 | 0.280 |
| Hispanic | High Income | % Overweight or Obese | -0.039 | 0.722 | 0.722 | 0.492 |
| Hispanic | High Education | Fresh F&V Consumption | 8.859 | 0.575 | 0.626 | < 0.001 |
| Hispanic | High Education | Fast Food Consumption | -11.902 | 0.385 | 0.339 | < 0.001 |
| Hispanic | High Education | Soda Consumption | -8.530 | 0.034 | 0.031 | 0.061 |
| Hispanic | High Education | BMI | -5.949 | 29.738 | 27.969 | < 0.001 |
| Hispanic | High Education | % Overweight or Obese | -13.697 | 0.758 | 0.654 | < 0.001 |
| Hispanic | Low Fast Food | Fresh F&V Consumption | 1.501 | 0.562 | 0.570 | 0.083 |
| Hispanic | Low Fast Food | Fast Food Consumption | -5.920 | 0.423 | 0.398 | < 0.001 |
| Hispanic | Low Fast Food | Soda Consumption | -0.538 | 0.038 | 0.038 | 0.433 |
| Hispanic | Low Fast Food | BMI | -0.172 | 29.750 | 29.699 | 0.291 |
| Hispanic | Low Fast Food | % Overweight or Obese | -1.800 | 0.763 | 0.750 | 0.004 |
| Hispanic | High Grocery | Fresh F&V Consumption | 7.351 | 0.525 | 0.563 | < 0.001 |
| Hispanic | High Grocery | Fast Food Consumption | -7.212 | 0.443 | 0.411 | < 0.001 |
| Hispanic | High Grocery | Soda Consumption | -1.956 | 0.040 | 0.039 | 0.178 |
| Hispanic | High Grocery | BMI | -1.518 | 30.358 | 29.898 | < 0.001 |
| Hispanic | High Grocery | % Overweight or Obese | -3.492 | 0.791 | 0.763 | < 0.001 |
| White | High Income | Fresh F&V Consumption | 2.182 | 0.643 | 0.657 | < 0.001 |
| White | High Income | Fast Food Consumption | -5.063 | 0.359 | 0.341 | < 0.001 |
| White | High Income | Soda Consumption | -5.512 | 0.037 | 0.035 | < 0.001 |
| White | High Income | BMI | 0.486 | 27.759 | 27.894 | < 0.001 |
| White | High Income | % Overweight or Obese | 3.261 | 0.625 | 0.646 | < 0.001 |
| White | High Education | Fresh F&V Consumption | 9.690 | 0.600 | 0.658 | < 0.001 |
| White | High Education | Fast Food Consumption | -5.878 | 0.363 | 0.342 | < 0.001 |
| White | High Education | Soda Consumption | -4.346 | 0.036 | 0.035 | < 0.001 |
| White | High Education | BMI | -4.100 | 28.955 | 27.768 | < 0.001 |
| White | High Education | % Overweight or Obese | -11.105 | 0.717 | 0.638 | < 0.001 |
| White | Low Fast Food | Fresh F&V Consumption | 6.028 | 0.625 | 0.663 | < 0.001 |
| White | Low Fast Food | Fast Food Consumption | -6.611 | 0.359 | 0.335 | < 0.001 |
| White | Low Fast Food | Soda Consumption | -16.363 | 0.039 | 0.033 | < 0.001 |
| White | Low Fast Food | BMI | -0.644 | 28.147 | 27.966 | < 0.001 |
| White | Low Fast Food | % Overweight or Obese | -2.373 | 0.662 | 0.647 | < 0.001 |
| White | High Grocery | Fresh F&V Consumption | 1.655 | 0.634 | 0.644 | < 0.001 |
| White | High Grocery | Fast Food Consumption | -5.004 | 0.368 | 0.349 | < 0.001 |
| White | High Grocery | Soda Consumption | -5.319 | 0.038 | 0.036 | < 0.001 |
| White | High Grocery | BMI | -0.104 | 28.239 | 28.209 | 0.082 |
| White | High Grocery | % Overweight or Obese | -1.373 | 0.665 | 0.656 | < 0.001 |

**Table 5:** Effect sizes of all top/bottom quartiles matching experiments (Fig. 6).

| Treatment | Outcome | % Change | Trt. Mean | Ctrl. Mean | P-value (bootstrapping.) |
| --- | --- | --- | --- | --- | --- |
| High Income | Fresh F&V Consumption | 7.383 | 0.636 | 0.683 | < 0.001 |
| High Income | Fast Food Consumption | -9.366 | 0.345 | 0.312 | < 0.001 |
| High Income | Soda Consumption | -7.064 | 0.032 | 0.029 | < 0.001 |
| High Income | BMI | 0.689 | 27.175 | 27.362 | < 0.001 |
| High Income | % Overweight or Obese | 2.904 | 0.596 | 0.613 | < 0.001 |
| High Education | Fresh F&V Consumption | 13.977 | 0.582 | 0.664 | < 0.001 |
| High Education | Fast Food Consumption | -10.323 | 0.382 | 0.342 | < 0.001 |
| High Education | Soda Consumption | -12.293 | 0.039 | 0.034 | < 0.001 |
| High Education | BMI | -6.641 | 29.510 | 27.550 | < 0.001 |
| High Education | % Overweight or Obese | -15.904 | 0.739 | 0.621 | < 0.001 |
| High Grocery | Fresh F&V Consumption | 5.664 | 0.614 | 0.649 | < 0.001 |
| High Grocery | Fast Food Consumption | -11.619 | 0.379 | 0.335 | < 0.001 |
| High Grocery | Soda Consumption | -14.903 | 0.038 | 0.032 | < 0.001 |
| High Grocery | BMI | -0.646 | 28.564 | 28.379 | < 0.001 |
| High Grocery | % Overweight or Obese | -2.613 | 0.683 | 0.665 | < 0.001 |
| Low Fast Food | Fresh F&V Consumption | 8.893 | 0.613 | 0.668 | < 0.001 |
| Low Fast Food | Fast Food Consumption | -11.542 | 0.371 | 0.328 | < 0.001 |
| Low Fast Food | Soda Consumption | -30.282 | 0.042 | 0.029 | < 0.001 |
| Low Fast Food | BMI | -0.153 | 28.161 | 28.118 | 0.098 |
| Low Fast Food | % Overweight or Obese | -0.384 | 0.656 | 0.654 | 0.140 |

**Table 6:**
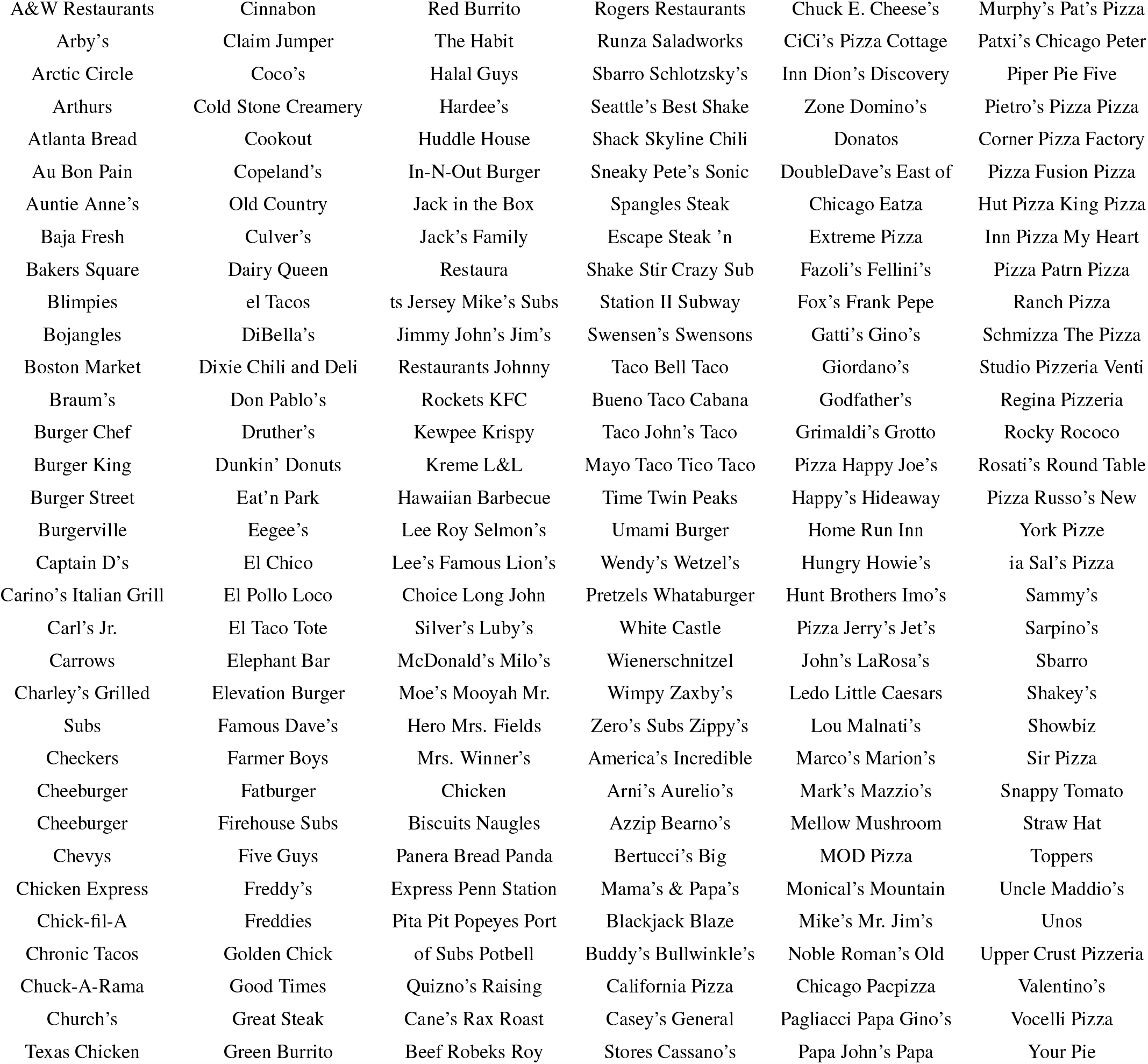
USA fast food restaurants table used to classify participant food entries as fast food^88^. A list of popular pizza chains from the USA was appended to the list^89^.

**Table 7:** USA soda list used to classify participant food entries as sugary sodas. The list was constructed using a list of America’s best-selling brands of Soda^90^, in addition to the generic terms such “Root Beer” and “Coca”.

|  |  |  |
| --- | --- | --- |
| Pepsi | Dr Pepper | Root Beer |
| Coca Cola | Sprite | Coke |
| Mountain Dew | Fanta | Coca |

#### Data Validation

We find that our study population has significant overlap with the U.S. national population (Table 1) but is skewed towards women and higher income. We demonstrate that food consumption measured based on this population are highly correlated with state-of-the-art measures (Figure 3, Section 2.3). Smartphone apps such as MyFitnessPal feature large databases with nutritional information and can be used to track one’s diet over time. Previous studies have compared app-reported diet measures to traditional measures including 24 hour dietary recalls and food composition tables. These studies found that both measures tend to be highly correlated^80, 81^, but that app-reported measures tend to underestimate certain macro- and micronutrients^80, 81^, especially in populations that were previously unfamiliar with the smartphone applications^82^. In contrast, this study leverages a convenience sample of existing participants of the smartphone app MyFitnessPal. Yelp data has been used in measures of food environment^83^ and a study in Detroit found Yelp data to be more accurate than commercially-available databases such as Reference USA^84^. This study uses a combination of MFP data to capture food consumption, Yelp, and USDA data to capture food environment, and Census data to capture basic demographics. As a preliminary, basic test, we investigated correlations between the Mexican food consumption, the fraction of Mexican restaurants, and the fraction of Hispanics in the population, on a zip code level. We found that Mexican food consumption (entries labeled as Mexican food by a proprietary MFP classifier, logged per participant, per day) was correlated with the fraction of Mexican restaurants (R=0.72; < 10^−4^) and the fraction of Hispanics in the population (R=0.54; *P* < 10^−4^). Further, the fraction of Mexican restaurants was correlated with the fraction of Hispanics in the population as well (R=0.51; *P* < 10^−4^).

#### Details on reproducing state-of-the-art measures using population-scale digital food logs

We used the latest survey data from BRFSS^2, 3^ available at the county-level. Specifically, we used variables FV5SRV from BRFSS 2011 representing the the faction of people eating five or more servings of fresh fruit and vegetables^2^, and BMI5 from BRFSS 2012 representing body mass index^3^. Comparing to BRFSS on a county level, the average number of F&V logged per day (MFP) was correlated with the fraction of respondents that report consumptions of at least five servings of F&V per day (Figure 3a, R=0.63, *p* < 10^−5^). Further, average county-level BMI was strongly correlated as well (Figure 3b, R=0.78, *p* < 10^−5^). We further compared to published results by the USDA^4^, which used data from the 2010 Nielsen Homescan Panel Survey that captured household food purchases for in-home consumption (but did not capture restaurants and fast food purchases). We attempted to reproduce published findings on the differences in low-income, low-access communities (food deserts) compared to non-low-income, non-low-access communities^4^ across categories of fruit, vegetable, sweets, red meat, fish/poultry, milk products, diet drinks, and non-diet drinks (Table 4 in Rahkovsky and Snyder^4^). We used proprietary MFP classifiers to categorize foods logged into these categories. We found that our app-based food logs were very highly correlated with previously published results (Figure 3c, R=0.88, *p* < 0.01) and that the absolute differences between food deserts and non-food deserts were stronger in the MFP data compared to Nielsen purchase data. In total, these results demonstrate convergent validity. Specifically, our results suggest that population-scale digital food logs can reproduce the basic dynamics of traditional, state-of-the-art measures, and they can do so at massive scale and comparatively low cost.

#### Details on Matching Approach

Specifically, we use a one-to-one Genetic Matching approach^79^, with replacement, and use the mean of the Standardized Mean Difference (SMD) between treatment and control groups, across all matched variables, as the Genetic Matching balance metric in order to maximize balance (overlap) between the treated and the control units. Some definitions of SMD use the standard variation in the overall population before matching^29^. However, we choose the standard deviation in the control group post-matching, which typically is much smaller and therefore gives more conservative estimates of balance between treated and control units^85^.

After matching, we evaluated the quality of balance between the treated and the control units by the Standardized Mean Difference across each of the variables that were controlled for and included in the matching process. A good balance between treated and control groups was defined as a Standardized Mean Difference (SMD) of less than 0.25 standard deviations^30^ across each variable. By default, we do not enforce a caliper in order to minimize bias in matching process, although in rare cases in which a good balance was not achieved, a caliper was enforced, starting at 2.5 standard deviations between matched and controlled units, and decreased by 0.1 until the matched and control groups had a SMD smaller than 0.25 across all matched variables.

For the vast majority of matching experiments the SMD across all matched variables was well below 0.25, with a mean of 0.040 and median of 0.016 for the four overall population matching experiments. The SMD for the race-majority zipcode experiments was slightly higher, but still very significantly below 0.25 across all 12 experiments, with a mean of 0.055 and median of 0.036. Thus, no caliper was necessary to ensure a good balance, with the exception of one out of the 12 of sub-population experiments (White, High Education). Detailed balancing statistics for each of the matches are available in the Appendix (Tables 8a-30b), as well as a supplementary matching experiment in which a top/bottom quartile split was used instead of a median split (Figure 6).

**Table 8:** Summary of High Income (MedianFamilyIncome > Median) matching experiment

(a) Sample sizes
|  | Control | Treated |
| --- | --- | --- |
| All | 4911 | 4911 |
| Matched | 1358 | 4911 |
| Unmatched | 3553 | 0 |
| Discarded | 0 | 0 |

(b) Summary of balance for matched data
|  | Means Treated | Means Control | SD Control | Mean Diff | Std. Mean Difference |
| --- | --- | --- | --- | --- | --- |
| distance | 0.75 | 0.75 | 0.26 | 0.00 | 0.01 |
| Education (% Without College Degree) | 0.56 | 0.57 | 0.15 | -0.01 | -0.06 |
| Grocery Distance (USDA lapophalfshare) | 0.74 | 0.74 | 0.21 | 0.00 | 0.00 |
| Yelp Fast Food % | 0.06 | 0.06 | 0.06 | 0.00 | -0.01 |
| Median Family Income | 97244.90 | 58991.91 | 8680.39 | 38253.00 | 4.41 |

**Table 9:** Summary of High Grocery (Grocery Access > Median) matching experiment

(a) Sample sizes
|  | Control | Treated |
| --- | --- | --- |
| All | 4911 | 4911 |
| Matched | 2219 | 4911 |
| Unmatched | 2692 | 0 |
| Discarded | 0 | 0 |

(b) Summary of balance for matched data
|  | Means Treated | Means Control | SD Control | Mean Diff | Std. Mean Difference |
| --- | --- | --- | --- | --- | --- |
| distance | 0.59 | 0.58 | 0.19 | 0.00 | 0.01 |
| Median Family Income | 76817.91 | 77021.36 | 30133.94 | -203.45 | -0.01 |
| Education (% Without College Degree) | 0.64 | 0.64 | 0.18 | 0.00 | -0.01 |
| Yelp Fast Food % | 0.06 | 0.06 | 0.06 | 0.00 | 0.00 |
| Grocery Distance (USDA lapophalfshare) | 0.57 | 0.88 | 0.06 | -0.31 | -5.62 |

**Table 10:** Summary of High Education (% College Degrees > Median) matching experiment

(a) Sample sizes
|  | Control | Treated |
| --- | --- | --- |
| All | 4911 | 4911 |
| Matched | 1481 | 4911 |
| Unmatched | 3430 | 0 |
| Discarded | 0 | 0 |

(b) Summary of balance for matched data
|  | Means Treated | Means Control | SD Control | Mean Diff | Std. Mean Difference |
| --- | --- | --- | --- | --- | --- |
| distance | 0.73 | 0.73 | 0.27 | 0.00 | 0.01 |
| Median Family Income | 93203.36 | 88493.86 | 20530.36 | 4709.51 | 0.23 |
| Grocery Distance (USDA lapophalfshare) | 0.71 | 0.72 | 0.22 | -0.01 | -0.05 |
| Yelp Fast Food % | 0.06 | 0.06 | 0.06 | 0.00 | 0.02 |
| Education (% Without College Degree) | 0.53 | 0.75 | 0.05 | -0.22 | -4.79 |

**Table 11:** Summary of Low Fast Food (% Yelp Fast Food < Median) matching experiment

(a) Sample sizes
|  | Control | Treated |
| --- | --- | --- |
| All | 4911 | 4911 |
| Matched | 1910 | 4911 |
| Unmatched | 3001 | 0 |
| Discarded | 0 | 0 |

(b) Summary of balance for matched data
|  | Means Treated | Means Control | SD Control | Mean Diff | Std. Mean Difference |
| --- | --- | --- | --- | --- | --- |
| distance | 0.65 | 0.65 | 0.24 | 0.00 | 0.01 |
| Median Family Income | 86942.10 | 86597.30 | 31088.00 | 344.80 | 0.01 |
| Education (% Without College Degree) | 0.60 | 0.61 | 0.18 | 0.00 | -0.02 |
| Grocery Distance (USDA lapophalfshare) | 0.66 | 0.67 | 0.23 | -0.01 | -0.05 |
| Yelp Fast Food % | 0.03 | 0.11 | 0.06 | -0.08 | -1.35 |

**Table 12:** Summary of High Income (MedianFamilyIncome > 75th Percentile) matching experiment

(a) Sample sizes
|  | Control | Treated |
| --- | --- | --- |
| All | 2319 | 2321 |
| Matched | 215 | 2321 |
| Unmatched | 2104 | 0 |
| Discarded | 0 | 0 |

(b) Summary of balance for matched data
|  | Means Treated | Means Control | SD Control | Mean Diff | Std. Mean Difference |
| --- | --- | --- | --- | --- | --- |
| distance | 0.91 | 0.91 | 0.18 | 0.00 | 0.01 |
| Education (% Without College Degree) | 0.47 | 0.48 | 0.13 | -0.02 | -0.11 |
| Grocery Distance (USDA lapophalfshare) | 0.71 | 0.70 | 0.22 | 0.01 | 0.04 |
| Yelp Fast Food % | 0.04 | 0.04 | 0.04 | 0.00 | -0.06 |
| Median Family Income | 115443.80 | 48357.34 | 6021.64 | 67086.46 | 11.14 |

**Table 13:** Summary of High Grocery (Grocery Access > 75th Percentile) matching experiment

(a) Sample sizes
|  | Control | Treated |
| --- | --- | --- |
| All | 2321 | 2319 |
| Matched | 743 | 2319 |
| Unmatched | 1578 | 0 |
| Discarded | 0 | 0 |

(b) Summary of balance for matched data
|  | Means Treated | Means Control | SD Control | Mean Diff | Std. Mean Difference |
| --- | --- | --- | --- | --- | --- |
| distance | 0.69 | 0.68 | 0.23 | 0.01 | 0.03 |
| Median Family Income | 77354.53 | 77809.00 | 31756.28 | -454.47 | -0.01 |
| Education (% Without College Degree) | 0.62 | 0.62 | 0.19 | 0.00 | -0.01 |
| Yelp Fast Food % | 0.04 | 0.04 | 0.04 | 0.00 | -0.01 |
| Grocery Distance (USDA lapophalfshare) | 0.41 | 0.94 | 0.04 | -0.52 | -13.78 |

**Table 14:**
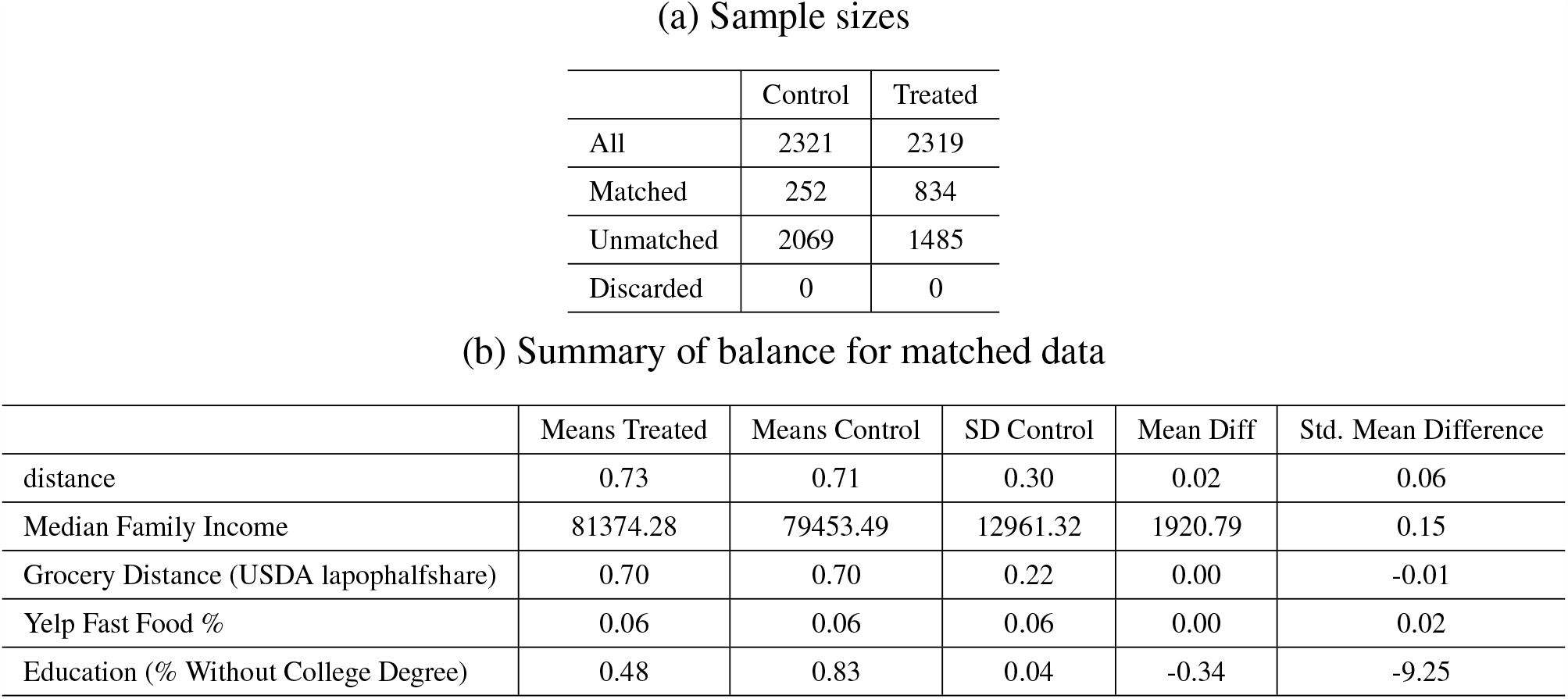
Summary of High Education (% College Degrees > 75th Percentile) matching experiment. **Note:** Treatment samples dropped due to 0.3 STD caliper used to ensure 0.25 SMD balancing constraint.

**Table 15:** Summary of Low Fast Food (% Yelp Fast Food < 25th Percentile) matching experiment. **Note:** Treatment samples dropped due to 1.5 STD caliper used to ensure 0.25 SMD balancing constraint.

|  | Control | Treated |
| --- | --- | --- |
| All | 2322 | 2319 |
| Matched | 504 | 2166 |
| Unmatched | 1818 | 153 |
| Discarded | 0 | 0 |

(b) Summary of balance for matched data
|  | Means Treated | Means Control | SD Control | Mean Diff | Std. Mean Difference |
| --- | --- | --- | --- | --- | --- |
| distance | 0.78 | 0.77 | 0.27 | 0.01 | 0.04 |
| Median Family Income | 88631.94 | 83632.13 | 20794.81 | 4999.81 | 0.24 |
| Education (% Without College Degree) | 0.59 | 0.59 | 0.15 | 0.00 | -0.02 |
| Grocery Distance (USDA lapophalfshare) | 0.63 | 0.66 | 0.23 | -0.03 | -0.14 |
| Yelp Fast Food % | 0.02 | 0.18 | 0.05 | -0.16 | -3.23 |

**Table 16:** Summary of Low Countertop Installation Services (% Yelp Countertop Installers < Median) matching experiment

(a) Sample sizes
|  | Control | Treated |
| --- | --- | --- |
| All | 4913 | 4909 |
| Matched | 2829 | 4909 |
| Unmatched | 2084 | 0 |
| Discarded | 0 | 0 |

(b) Summary of balance for matched data
|  | Means Treated | Means Control | SD Control | Mean Diff | Std. Mean Difference |
| --- | --- | --- | --- | --- | --- |
| distance | 0.50 | 0.50 | 0.05 | 0.00 | 0.01 |
| Median Family Income | 77255.32 | 77443.29 | 28361.82 | -187.96 | -0.01 |
| Education (% Without College Degree) | 0.65 | 0.66 | 0.18 | 0.00 | 0.00 |
| Grocery Distance (USDA lapophalfshare) | 0.72 | 0.72 | 0.24 | 0.00 | 0.00 |
| Yelp Fast Food % | 0.09 | 0.09 | 0.08 | 0.00 | 0.01 |
| Yelp Countertop Installers % | 0.00 | 0.00 | 0.00 | 0.00 | -1.61 |

**Table 17:** Summary of Low Electronics Stores (% Yelp Electronics Stores < Median) matching experiment

(a) Sample sizes
|  | Control | Treated |
| --- | --- | --- |
| All | 4912 | 4910 |
| Matched | 2741 | 4910 |
| Unmatched | 2171 | 0 |
| Discarded | 0 | 0 |

(b) Summary of balance for matched data
|  | Means Treated | Means Control | SD Control | Mean Diff | Std. Mean Difference |
| --- | --- | --- | --- | --- | --- |
| distance | 0.52 | 0.52 | 0.09 | 0.00 | 0.00 |
| Median Family Income | 74567.18 | 74306.84 | 25820.76 | 260.34 | 0.01 |
| Education (% Without College Degree) | 0.68 | 0.68 | 0.16 | 0.00 | 0.00 |
| Grocery Distance (USDA lapophalfshare) | 0.71 | 0.71 | 0.22 | 0.00 | -0.01 |
| Yelp Fast Food % | 0.08 | 0.08 | 0.07 | 0.00 | 0.00 |
| Yelp Electronics Stores % | 0.00 | 0.00 | 0.00 | 0.00 | -1.59 |

**Table 18:** Summary of Low Waterproofing Services (% Yelp Waterproofing Services < Median) matching experiment

(a) Sample sizes
|  | Control | Treated |
| --- | --- | --- |
| All | 4913 | 4909 |
| Matched | 2876 | 4909 |
| Unmatched | 2037 | 0 |
| Discarded | 0 | 0 |

(b) Summary of balance for matched data
|  | Means Treated | Means Control | SD Control | Mean Diff | Std. Mean Difference |
| --- | --- | --- | --- | --- | --- |
| distance | 0.50 | 0.50 | 0.02 | 0.00 | 0.00 |
| Median Family Income | 76371.54 | 76438.89 | 28071.29 | -67.35 | 0.00 |
| Education (% Without College Degree) | 0.66 | 0.66 | 0.17 | 0.00 | 0.00 |
| Grocery Distance (USDA lapophalfshare) | 0.74 | 0.74 | 0.21 | 0.00 | -0.01 |
| Yelp Fast Food % | 0.09 | 0.09 | 0.08 | 0.00 | 0.01 |
| Yelp Waterproofing Services % | 0.00 | 0.00 | 0.00 | 0.00 | -1.48 |

**Table 19:** Summary of Black-majority Zip Code High Income (MedianFamilyIncome > Median) matching experiment

(a) Sample sizes
|  | Control | Treated |
| --- | --- | --- |
| All | 317 | 42 |
| Matched | 30 | 42 |
| Unmatched | 287 | 0 |
| Discarded | 0 | 0 |

(b) Summary of balance for matched data
|  | Means Treated | Means Control | SD Control | Mean Diff | Std. Mean Difference |
| --- | --- | --- | --- | --- | --- |
| distance | 0.36 | 0.35 | 0.25 | 0.02 | 0.07 |
| Education (% Without College Degree) | 0.65 | 0.65 | 0.09 | -0.01 | -0.08 |
| Grocery Distance (USDA lapophalfshare) | 0.68 | 0.67 | 0.22 | 0.01 | 0.02 |
| Yelp Fast Food % | 0.04 | 0.03 | 0.03 | 0.00 | 0.09 |
| Median Family Income | 88944.14 | 59779.41 | 8039.94 | 29164.73 | 3.63 |

**Table 20:**
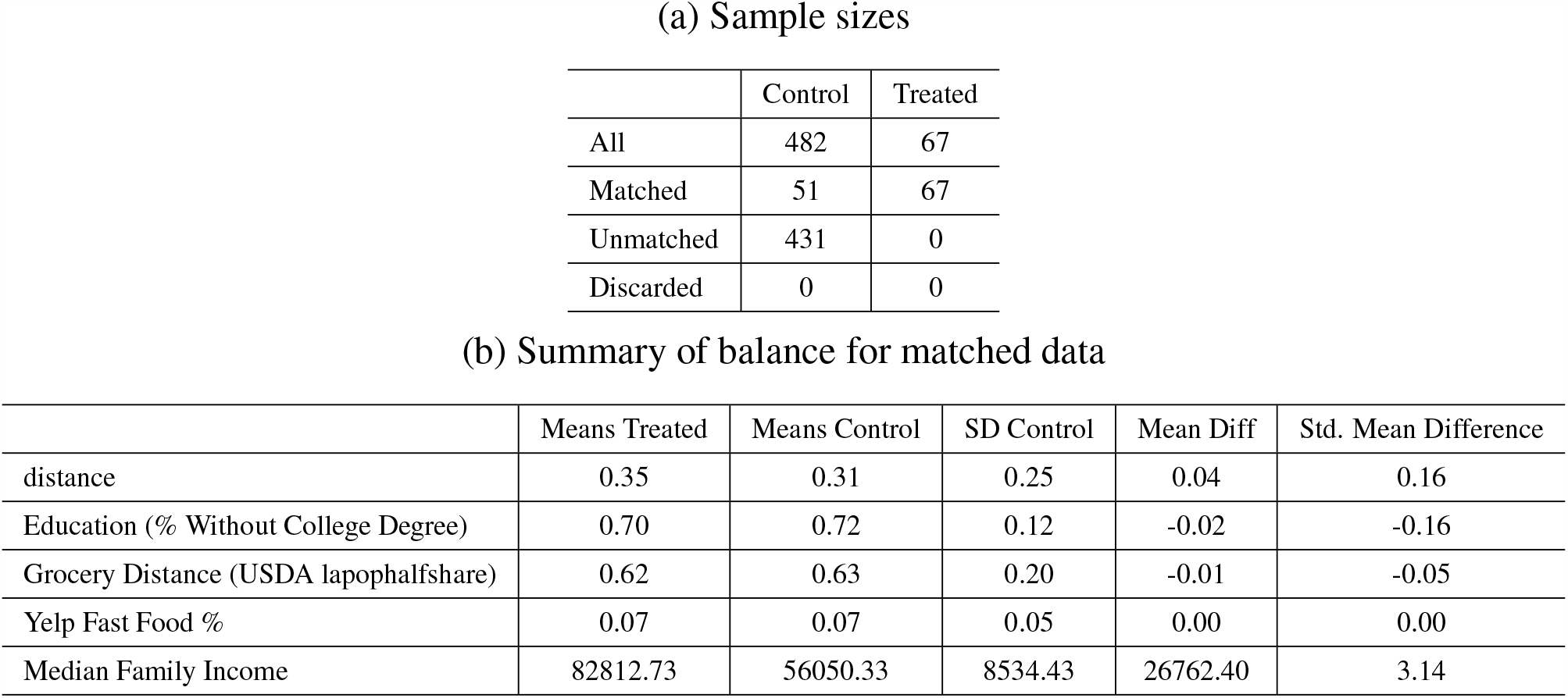
Summary of Hispanic-majority Zip Code High Income (MedianFamilyIncome > Median) matching experiment

**Table 21:** Summary of White-majority Zip Code High Income (MedianFamilyIncome > Median) matching experiment

(a) Sample sizes
|  | Control | Treated |
| --- | --- | --- |
| All | 3421 | 4277 |
| Matched | 1023 | 4277 |
| Unmatched | 2398 | 0 |
| Discarded | 0 | 0 |

(b) Summary of balance for matched data
|  | Means Treated | Means Control | SD Control | Mean Diff | Std. Mean Difference |
| --- | --- | --- | --- | --- | --- |
| distance | 0.78 | 0.78 | 0.25 | 0.00 | 0.01 |
| Education (% Without College Degree) | 0.55 | 0.56 | 0.15 | -0.01 | -0.07 |
| Grocery Distance (USDA lapophalfshare) | 0.76 | 0.76 | 0.19 | 0.00 | -0.01 |
| Yelp Fast Food % | 0.06 | 0.06 | 0.06 | 0.00 | -0.01 |
| Median Family Income | 98014.79 | 59878.82 | 8719.97 | 38135.97 | 4.37 |

**Table 22:** Summary of Black-majority Zip Code High Grocery (Grocery Access > Median) matching experiment

(a) Sample sizes
|  | Control | Treated |
| --- | --- | --- |
| All | 100 | 259 |
| Matched | 65 | 259 |
| Unmatched | 35 | 0 |
| Discarded | 0 | 0 |

(b) Summary of balance for matched data
|  | Means Treated | Means Control | SD Control | Mean Diff | Std. Mean Difference |
| --- | --- | --- | --- | --- | --- |
| distance | 0.78 | 0.77 | 0.15 | 0.01 | 0.08 |
| Median Family Income | 51410.82 | 51925.49 | 14357.87 | -514.66 | -0.04 |
| Education (% Without College Degree) | 0.76 | 0.77 | 0.08 | 0.00 | -0.03 |
| Yelp Fast Food % | 0.05 | 0.05 | 0.05 | 0.00 | -0.08 |
| Grocery Distance (USDA lapophalfshare) | 0.53 | 0.86 | 0.04 | -0.33 | -9.43 |

**Table 23:** Summary of Hispanic-majority Zip Code High Grocery (Grocery Access > Median) matching experiment

(a) Sample sizes
|  | Control | Treated |
| --- | --- | --- |
| All | 78 | 471 |
| Matched | 66 | 471 |
| Unmatched | 12 | 0 |
| Discarded | 0 | 0 |

(b) Summary of balance for matched data
|  | Means Treated | Means Control | SD Control | Mean Diff | Std. Mean Difference |
| --- | --- | --- | --- | --- | --- |
| distance | 0.87 | 0.87 | 0.09 | 0.00 | 0.01 |
| Median Family Income | 52218.86 | 53068.49 | 13918.72 | -849.63 | -0.06 |
| Education (% Without College Degree) | 0.82 | 0.83 | 0.08 | -0.01 | -0.12 |
| Yelp Fast Food % | 0.06 | 0.06 | 0.05 | 0.00 | 0.01 |
| Grocery Distance (USDA lapophalfshare) | 0.47 | 0.88 | 0.05 | -0.41 | -7.86 |

**Table 24:** Summary of White-majority Zip Code High Grocery (Grocery Access > Median) matching experiment

(a) Sample sizes
|  | Control | Treated |
| --- | --- | --- |
| All | 4494 | 3204 |
| Matched | 1741 | 3204 |
| Unmatched | 2753 | 0 |
| Discarded | 0 | 0 |

(b) Summary of balance for matched data
|  | Means Treated | Means Control | SD Control | Mean Diff | Std. Mean Difference |
| --- | --- | --- | --- | --- | --- |
| distance | 0.51 | 0.50 | 0.19 | 0.00 | 0.02 |
| Median Family Income | 84030.48 | 84275.98 | 31002.90 | -245.50 | -0.01 |
| Education (% Without College Degree) | 0.59 | 0.59 | 0.18 | 0.00 | -0.01 |
| Yelp Fast Food % | 0.07 | 0.07 | 0.07 | 0.00 | 0.00 |
| Grocery Distance (USDA lapophalfshare) | 0.62 | 0.89 | 0.05 | -0.27 | -4.92 |

**Table 25:** Summary of Black-majority Zip Code High Education (% College Degrees > Median) matching experiment

(a) Sample sizes
|  | Control | Treated |
| --- | --- | --- |
| All | 285 | 74 |
| Matched | 48 | 74 |
| Unmatched | 237 | 0 |
| Discarded | 0 | 0 |

(b) Summary of balance for matched data
|  | Means Treated | Means Control | SD Control | Mean Diff | Std. Mean Difference |
| --- | --- | --- | --- | --- | --- |
| distance | 0.46 | 0.45 | 0.29 | 0.01 | 0.02 |
| Median Family Income | 70932.69 | 71048.45 | 22123.91 | -115.77 | -0.01 |
| Grocery Distance (USDA lapophalfshare) | 0.59 | 0.60 | 0.28 | -0.01 | -0.04 |
| Yelp Fast Food % | 0.04 | 0.04 | 0.04 | 0.00 | 0.00 |
| Education (% Without College Degree) | 0.62 | 0.77 | 0.04 | -0.15 | -3.39 |

**Table 26:**
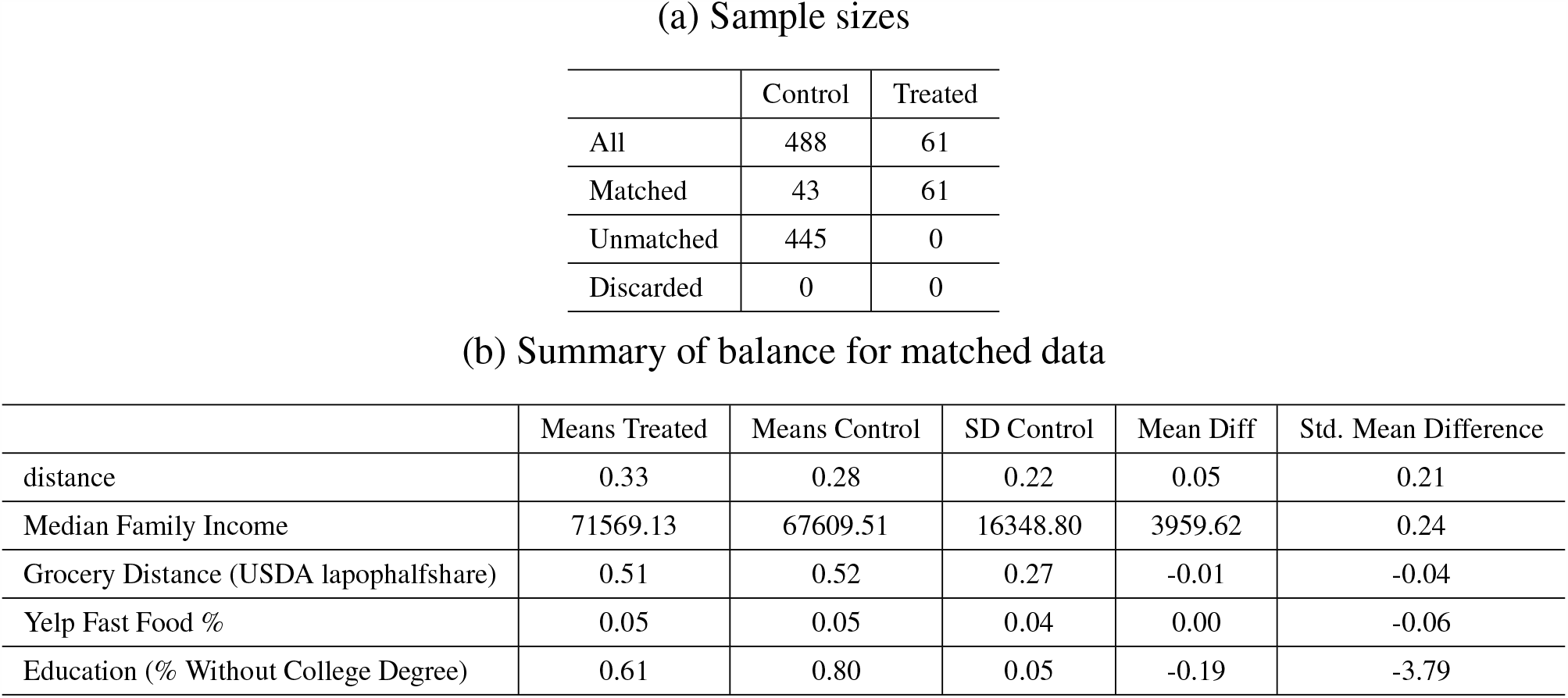
Summary of Hispanic-majority Zip Code High Education (% College Degrees > Median) matching experiment

**Table 27:** Summary of White-majority Zip Code High Education (% College Degrees > Median) matching experiment. **Note:** Treatment samples dropped due to 2.1 STD caliper used to ensure 0.25 SMD balancing constraint.

|  | Control | Treated |
| --- | --- | --- |
| All | 3491 | 4207 |
| Matched | 1114 | 4102 |
| Unmatched | 2377 | 105 |
| Discarded | 0 | 0 |

(b) Summary of balance for matched data
|  | Means Treated | Means Control | SD Control | Mean Diff | Std. Mean Difference |
| --- | --- | --- | --- | --- | --- |
| distance | 0.75 | 0.75 | 0.26 | 0.00 | 0.01 |
| Median Family Income | 92654.95 | 88357.13 | 17926.24 | 4297.82 | 0.24 |
| Grocery Distance (USDA lapophalfshare) | 0.74 | 0.76 | 0.17 | -0.01 | -0.07 |
| Yelp Fast Food % | 0.07 | 0.07 | 0.06 | 0.00 | 0.00 |
| Education (% Without College Degree) | 0.53 | 0.75 | 0.05 | -0.22 | -4.59 |

**Table 28:**
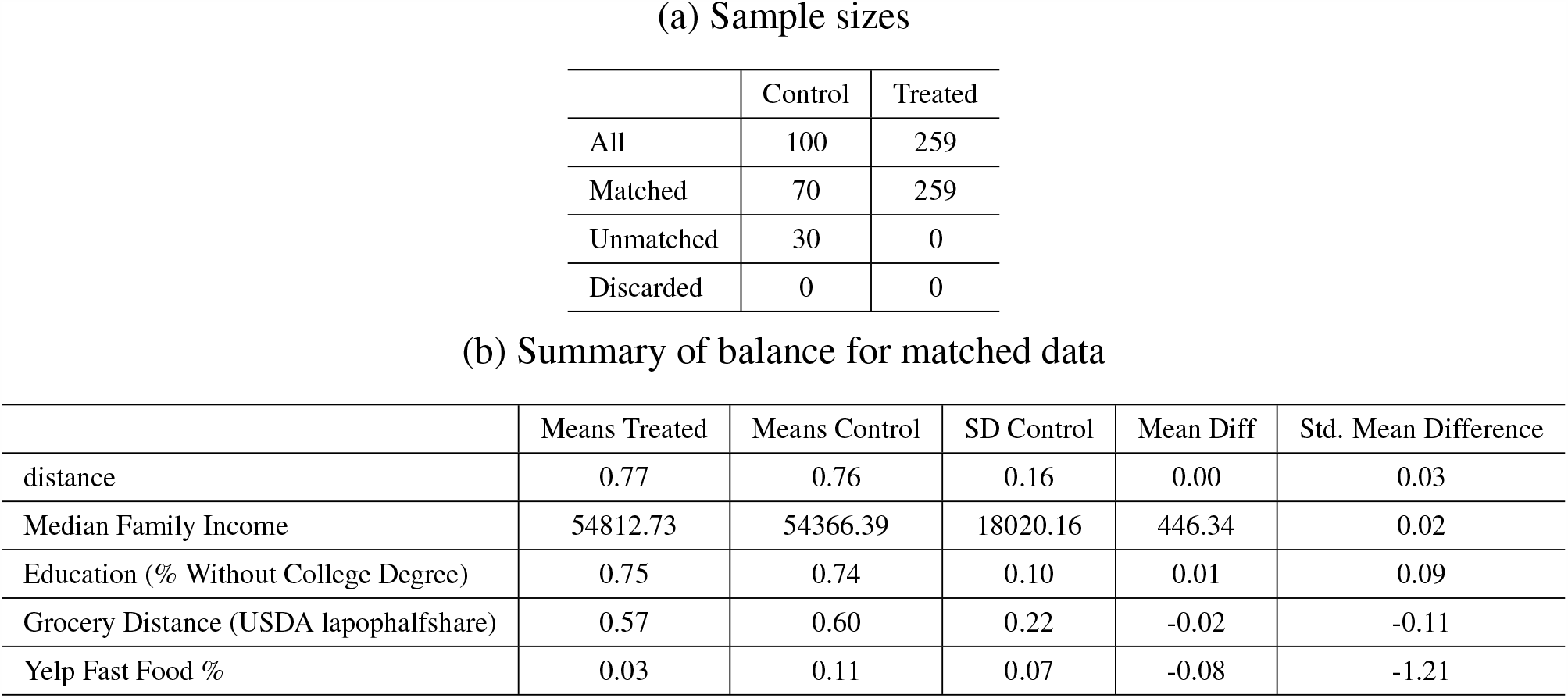
Summary of Black-majority Zip Code Low Fast Food (% Yelp Fast Food < Median) matching experiment

(a) Sample sizes
|  | Control | Treated |
| --- | --- | --- |
| All | 100 | 259 |
| Matched | 70 | 259 |
| Unmatched | 30 | 0 |
| Discarded | 0 | 0 |

(b) Summary of balance for matched data
|  | Means Treated | Means Control | SD Control | Mean Diff | Std. Mean Difference |
| --- | --- | --- | --- | --- | --- |
| distance | 0.77 | 0.76 | 0.16 | 0.00 | 0.03 |
| Median Family Income | 54812.73 | 54366.39 | 18020.16 | 446.34 | 0.02 |
| Education (% Without College Degree) | 0.75 | 0.74 | 0.10 | 0.01 | 0.09 |
| Grocery Distance (USDA lapophalfshare) | 0.57 | 0.60 | 0.22 | -0.02 | -0.11 |
| Yelp Fast Food % | 0.03 | 0.11 | 0.07 | -0.08 | -1.21 |

**Table 29:**
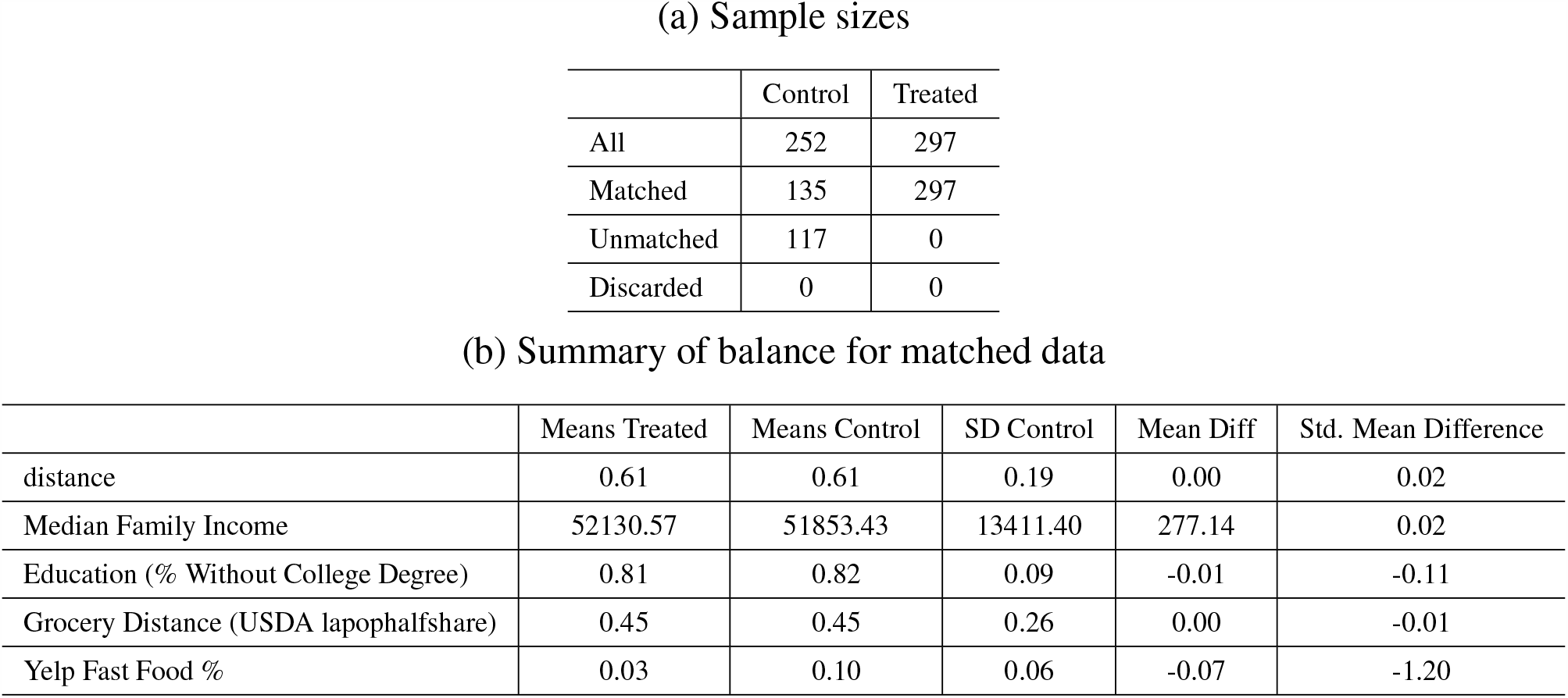
Summary of Hispanic-majority Zip Code Low Fast Food (% Yelp Fast Food < Median) matching experiment

(a) Sample sizes
|  | Control | Treated |
| --- | --- | --- |
| All | 252 | 297 |
| Matched | 135 | 297 |
| Unmatched | 117 | 0 |
| Discarded | 0 | 0 |

(b) Summary of balance for matched data
|  | Means Treated | Means Control | SD Control | Mean Diff | Std. Mean Difference |
| --- | --- | --- | --- | --- | --- |
| distance | 0.61 | 0.61 | 0.19 | 0.00 | 0.02 |
| Median Family Income | 52130.57 | 51853.43 | 13411.40 | 277.14 | 0.02 |
| Education (% Without College Degree) | 0.81 | 0.82 | 0.09 | -0.01 | -0.11 |
| Grocery Distance (USDA lapophalfshare) | 0.45 | 0.45 | 0.26 | 0.00 | -0.01 |
| Yelp Fast Food % | 0.03 | 0.10 | 0.06 | -0.07 | -1.20 |

**Table 30:** Summary of White-majority Zip Code Low Fast Food (% Yelp Fast Food < Median) matching experiment

(a) Sample sizes
|  | Control | Treated |
| --- | --- | --- |
| All | 4188 | 3510 |
| Matched | 1362 | 3510 |
| Unmatched | 2826 | 0 |
| Discarded | 0 | 0 |

(b) Summary of balance for matched data
|  | Means Treated | Means Control | SD Control | Mean Diff | Std. Mean Difference |
| --- | --- | --- | --- | --- | --- |
| distance | 0.65 | 0.65 | 0.26 | 0.00 | 0.01 |
| Median Family Income | 95359.35 | 94854.08 | 29365.91 | 505.27 | 0.02 |
| Education (% Without College Degree) | 0.56 | 0.57 | 0.17 | 0.00 | -0.01 |
| Grocery Distance (USDA lapophalfshare) | 0.71 | 0.72 | 0.20 | -0.01 | -0.05 |
| Yelp Fast Food % | 0.03 | 0.11 | 0.06 | -0.08 | -1.37 |

#### Details on obesity prevention related cost savings

It was estimated that the aggregate costs of obesity in the non-institutionalized adult population of the U.S. was as high as $315.8 billion in 2010, and the estimated extra annual medical care cost of an obese adult was $3,429 on average in 2013. The extra medical care costs are even higher for non-White populations ($4,086)^86^. The prevalence of overweight and obesity among US adults was 42.4% and about 107.6 million adults in 2017-2018^87^. A 13.1% decrease by implementing effective education programs and policies (*i*.*e*., based on our estimate of above/below median effect size, see Figure 4), could potentially lead to more than $48.3 billions of annual health care cost savings^35^. To produce this estimate we need to assume that the Average Treatment Effect on the Treated (ATT; estimated through our matching procedure) can be generalized to the overall U.S. population. According to the U.S. Department of Education, the 2019 president’s education budget for the entire U.S. was $64 billion^36^, which is significantly less than the aggregate costs of obesity. The national postsecondary education budget, which include important funding supporting upward mobility such as the federal Pell grant, work study and student loans, was $24.2 billion in 2020^36^. Based on our analysis, implementing effective education programs and policies could potentially lead to more than $48.3 billions of annual medical care cost savings in 2013 dollars. Taking inflation into account, this cost saving would be $53.6 billion in 2020, and could support 83.8% of the education budget and cover the entire postsecondary budget twice. Effective program and policy interventions could range from mandatory schooling policies, such as those promoting health and nutrition education at schools, to increasing educational quality, such as those aimed at promoting higher levels of education^37–39^. Similarly, having higher grocery access (2.4% decrease in overweight and obesity), and lower fast food access (1.5% decrease in overweight and obesity) could potentially lead to $9.9 billions and $6.1 billions of annual health care cost savings today respectively (based on our estimates in Figure 4).

